# Transcriptome profiling to identify blood biomarkers for peritoneal endometriosis

**DOI:** 10.64898/2025.12.23.25342915

**Authors:** Maja Pušić Novak, Angelika Vižintin, Tadeja Režen, Helena Ban Frangež, René Wenzl, Tea Lanišnik Rižner

## Abstract

**Context:** Peritoneal endometriosis (PE) remains challenging to diagnose, as it cannot be detected using standard imaging modalities and no clinically validated biomarkers are available.

**Objective:** To identify novel blood-based biomarkers for PE using whole blood transcriptomics combined with machine learning approaches.

**Design, Setting, and Patients:** This observational study enrolled 48 women undergoing laparoscopic surgery for endometriosis-related symptoms at tertiary referral centres in Slovenia and Austria. Patients were classified as having PE (n=20), peritoneal and ovarian endometriosis (PE and OE, n=8), or no endometriosis (controls, n=20). Patients were further stratified by menstrual phase (proliferative or secretory). Whole blood samples were collected preoperatively.

**Methods:** Whole-blood RNA sequencing was performed, and differentially expressed genes (DGEs) and transcripts (DTEs) were identified. Sequencing data were processed using a machine learning pipeline to select key features and develop support vector machine (SVM) classifiers for predicting endometriosis status.

**Results:** In the proliferative group, no DGEs and only two DTEs distinguished PE from controls. In contrast, in the secretory group, 1,035 DGEs and 922 DTEs were identified, with no overlap between menstrual phases. Enrichment analysis of secretory phase DGEs indicated their involvement in angiogenesis and immune-related pathways. Feature selection identified six transcripts that achieved the best SVM classification performance in distinguishing cases from controls across both menstrual phases (AUC = 0.92, sensitivity = 75%, specificity = 100%).

**Conclusion:** This study provides first evidence that integrating whole-blood transcriptomics with machine learning can identify potential blood-based biomarkers for PE and highlights the influence of menstrual cycle phase. These findings require validation in larger, independent cohort.

## INTRODUCTION

Endometriosis is a complex chronic disease affecting approximately 10 % of women in their reproductive age ^1^. It is an oestrogen-dependent, and chronic inflammatory condition associated with infertility and chronic pelvic pain ^2^. Histologically, endometriosis is characterised by the ectopic presence of endometrial stromal and epithelial cells, often accompanied by hemosiderin-containing macrophages ^3^. Endometriotic lesions are primarily found on the surface of the peritoneum (superficial peritoneal endometriosis, PE), on the ovaries (ovarian endometrioma, OE) or as nodules that penetrate more than 5 mm beneath the peritoneum (deep endometriosis, DE) ^3,4^.

Patients with endometriosis present with a variety of non-specific symptoms that often overlap with those of other gynaecological and gastrointestinal diseases, such as infertility, uterine fibroids, inflammatory bowel syndrome, or pelvic inflammatory disease ^5,6^. Additionally, approximately 20 – 25 % of patients with confirmed endometriosis are asymptomatic ^7,8^. Despite the high prevalence of this disease, endometriosis symptoms are often trivialised, discouraging patients from seeking medical help sooner ^9^. Consequently, there is a significant delay in diagnosis worldwide, ranging from 4 to 11 years after symptom onset ^5,10,11^.

Traditionally, laparoscopy, the surgical visualisation of endometriotic lesions, has been considered the gold standard for diagnosing endometriosis. However, this invasive and costly procedure carries surgical risks and can show variable results if not confirmed histologically ^12,13^. Studies have shown that two-thirds of women undergoing laparoscopy are not diagnosed with endometriosis, suggesting that many undergo unnecessary surgery ^14^. While OE and DE can be diagnosed with imaging modalities such as transvaginal ultrasound (TVUS) and magnetic resonance imaging (MRI), these techniques are not useful for diagnosing superficial PE, which accounts for approximately 80 % of all endometriosis cases ^2,15^. Furthermore, common non-pigmented endometriotic lesions present in patients with superficial PE may even be missed at laparoscopy ^16,17^. Recently, the European Society of Human Reproduction and Embryology (ESHRE) guidelines have recommended that the diagnosis of endometriosis should be considered in patients with related symptoms. Confirmation should be achieved with imaging, reserving laparoscopy for cases where imaging results are negative or where empirical treatment has been unsuccessful or is inappropriate. Clinicians are advised against using biomarkers for the diagnosis of endometriosis, as there is currently no reliable or clinically validated biomarker available for this condition ^18^. However, identification of accurate, reliable, and appropriately validated non-invasive biomarkers for endometriosis is needed to reduce current diagnostic delays and accelerate patient treatment ^19,20^.

To date, non-invasive biomarkers for endometriosis have been investigated in various biological samples, including peripheral blood, urine, menstrual blood, saliva, faeces and cervical mucus. Among these, peripheral blood, particularly serum and plasma, has been used most frequently ^21,22^. In contrast to serum and plasma, whole blood does not require separation into its constituent components, reducing potential inter-sample variability during processing and storage, and resulting in more reproducible results ^23–25^. Furthermore, whole blood collection is a standardized and routine procedure, making it ideal for rapid point-of-care tests ^24^. As blood perfuses all organs, it provides insights into human physiology and health, acting as a reporter for both systemic and localised diseases. Consequently, transcriptome analysis of blood can identify gene expression signatures and biomarkers for diagnosis, prognosis, and treatment monitoring ^26–28^.

So far, different molecules have been considered as potential biomarkers for endometriosis, including inflammatory markers, apoptosis and endothelial markers, glycoproteins, growth factors, oxidative stress markers, miRNAs, circRNAs and lncRNAs ^25,29,30^. To identify new biomarkers for endometriosis, studies use either hypothesis-driven approaches, or hypothesis generating - high-throughput (‘omics’) techniques, or a combination of both ^31^. Over the past 25 years, high-throughput screening technologies have been widely used to generate large-scale biological datasets to improve understanding, diagnosis and treatment of endometriosis ^32,33^. Additionally, different machine learning approaches are increasingly integrated into bioinformatics studies to analyse large datasets, such as patients clinical and lifestyle data, imaging data, and the expression of proteins, genes, metabolites and their combinations, to establish models for the diagnosis of endometriosis ^34^. However, many of these studies do not specify the subtype of endometriosis present among the enrolled patients but instead analyse all types of the disease as a single entity. As different endometriosis subtypes exhibit different pathophysiological characteristics ^35^, it is unlikely that to identify biomarkers that diagnose all types of endometriosis with high sensitivity and specificity. Furthermore, studies often lack a proper description of the modelling approach and are rarely validated in larger, independent cohorts ^34,36^.

In our previous studies, we searched for biomarkers of endometriosis in serum, plasma, and peritoneal fluid samples using different approaches, including targeted metabolomics ^37,38^ and proteomics ^39–42^. In the present study, our aim was to identify a panel of blood biomarker candidates for PE using whole genome transcriptomics combined with machine learning techniques.

## MATERIALS AND METHODS

### Study design and patient selection

This study was approved by the National Medical Ethics Committee of the Republic of Slovenia (approval no. 120-5412019-5), and the Republic of Austria (approval no. 545/2010). Patients were recruited prospectively between January 2020 and December 2022 at the University Medical Centre (UMC) Ljubljana, Slovenia, and between November 2019 and January 2020 at the Medical University of Vienna, Austria (Figure 1). Inclusion criteria were symptoms suggestive of endometriosis (chronic pelvic pain and/or infertility), reproductive age (18 to 39 years), and willingness to participate in the study. All recruited patients signed a written informed consent form upon inclusion. Exclusion criteria were pregnancy at the time of surgery, history or presence of malignant diseases and/or autoimmune diseases such as rheumatoid arthritis, inflammatory bowel disease, or autoimmune thyroid disease. Whole blood samples were collected from all patients. All women included underwent laparoscopic surgery and were characterized by the presence (cases) or absence (controls) of endometriosis after histological examination. Patients diagnosed with endometriosis were further classified according to the revised American Society of Reproductive Medicine scoring system (rASRM). Each patient also completed a questionnaire detailing general information about diet, lifestyle, smoking status, recreational habits, and ethnic origin. The attending physician completed an additional questionnaire covering clinical and gynaecological data, including use of oral contraception or hormonal therapy, medication use, regularity of menstrual cycle, phase of menstrual cycle at the time of surgery, and a surgical report with staging and scoring of endometriosis, previous gynaecological surgeries and presence of other pathologies.

**Figure 1.**
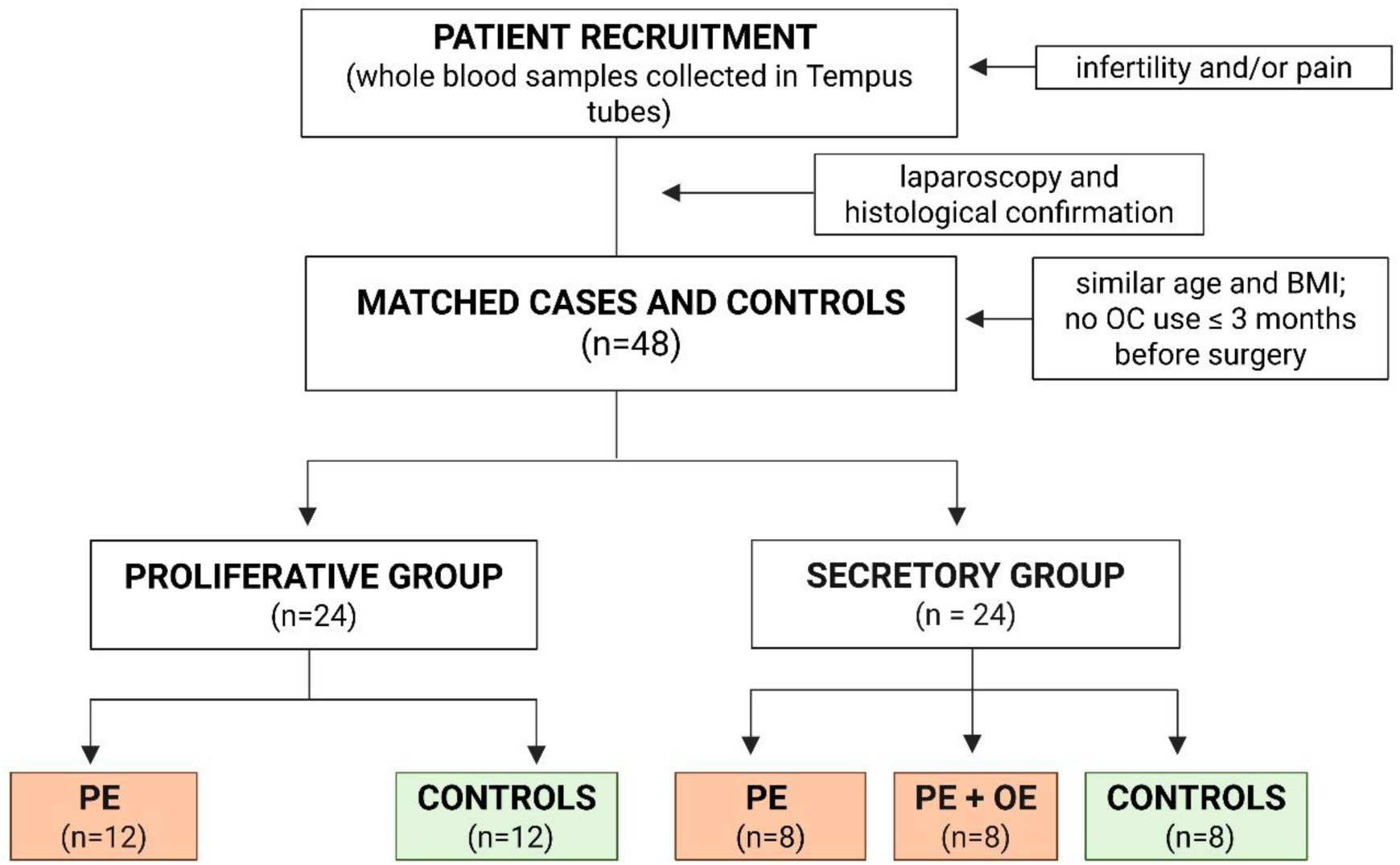
Flowchart of the patient selection. n-number, BMI – body mass index, PE - peritoneal endometriosis, OE – ovarian endometriosis, OC – oral contraception. Created with BioRender.com.

Case and control patients were carefully matched to ensure there were no significant differences in mean BMI or mean age. Furthermore, none of the participants had taken oral contraceptives in the three months preceding laparoscopy. Patients were divided into proliferative (n=24) and secretory (n=24) groups based on their menstrual cycle phase at the time of surgery. In the proliferative group, all cases (n=12) had PE, whereas no endometriosis was detected in the control patients (n=12). In the secretory group, cases (n=16) were divided into patients with PE (n=8) and those with both PE and OE (n=8). At the time of patient enrolment, there were insufficient numbers of patients in the secretory phase with PE only. Therefore, patients with combined PE and OE who matched the control criteria were also included. The absence of endometriosis was confirmed laparoscopically for controls (n=8). The clinical characteristics of patients in the discovery phase of the study are shown in Table 1.

**Table 1.**
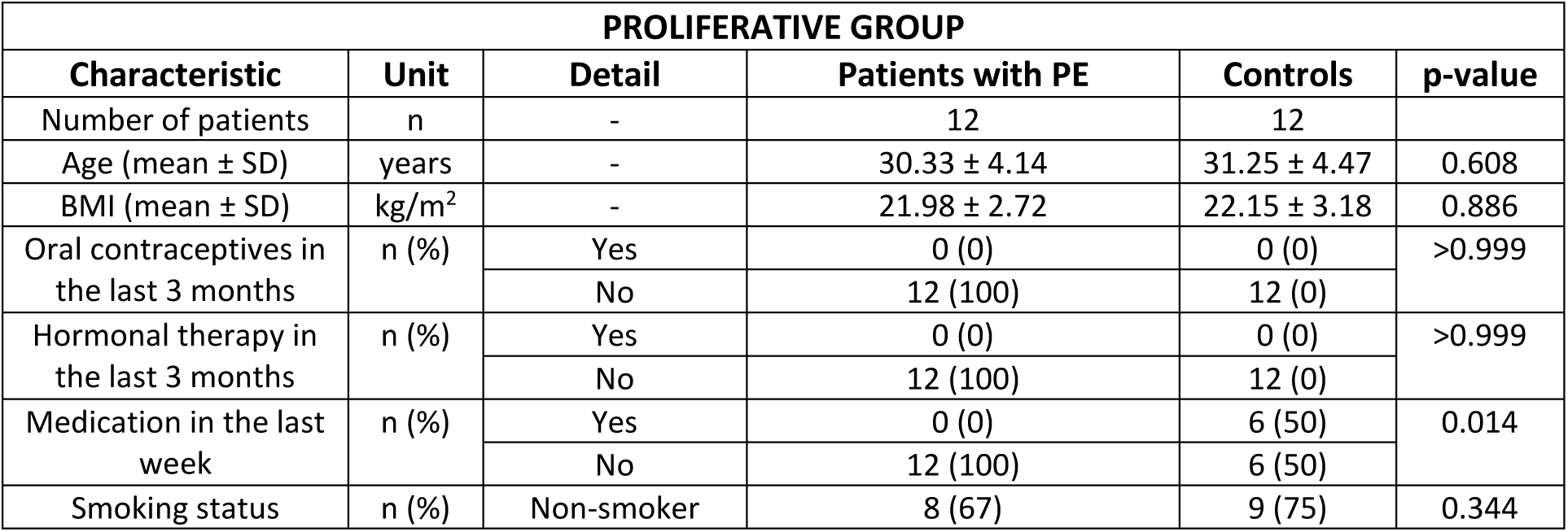
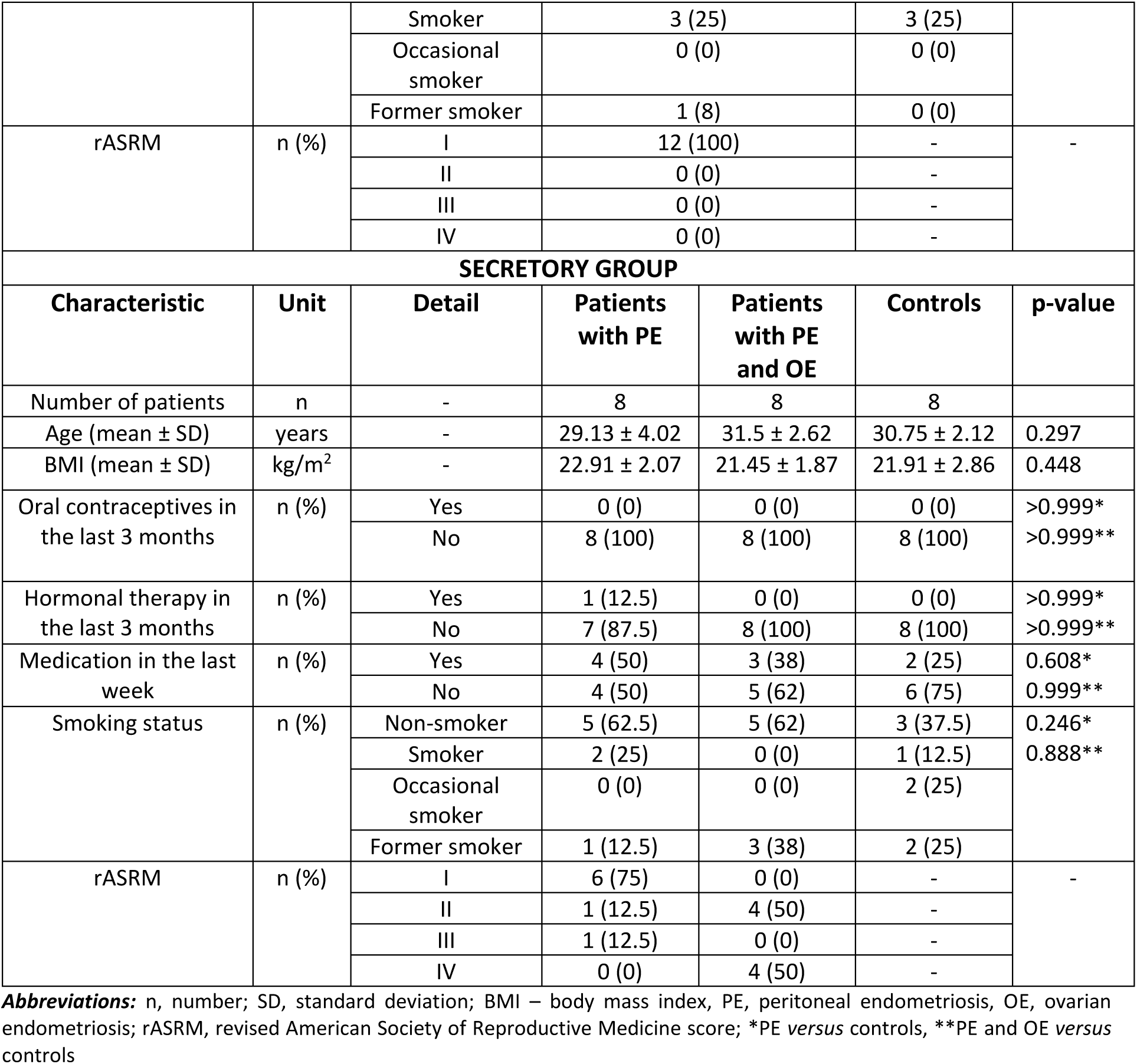
Clinical characteristics of patients included in the study.

| PROLIFERATIVE GROUP |  |  |  |  |  |
| --- | --- | --- | --- | --- | --- |
| Characteristic | Unit | Detail | Patients with PE | Controls | p-value |
| Number of patients | n | - | 12 | 12 |  |
| Age (mean ± SD) | years | - | 30.33 ± 4.14 | 31.25 ± 4.47 | 0.608 |
| BMI (mean ± SD) | kg/m<sup>2</sup> | - | 21.98 ± 2.72 | 22.15 ± 3.18 | 0.886 |
| Oral contraceptives in the last 3 months | n (%) | Yes | 0 (0) | 0 (0) | >0.999 |
|  |  | No | 12 (100) | 12 (0) |  |
| Hormonal therapy in the last 3 months | n (%) | Yes | 0 (0) | 0 (0) | >0.999 |
|  |  | No | 12 (100) | 12 (0) |  |
| Medication in the last week | n (%) | Yes | 0 (0) | 6 (50) | 0.014 |
|  |  | No | 12 (100) | 6 (50) |  |
| Smoking status | n (%) | Non-smoker | 8 (67) | 9 (75) | 0.344 |

|  |  |  |  |  |  |  |
| --- | --- | --- | --- | --- | --- | --- |
|  |  | Smoker | 3 (25) | 3 (25) |  |  |
|  |  | Occasional smoker | 0 (0) | 0 (0) |  |  |
|  |  | Former smoker | 1 (8) | 0 (0) |  |  |
| rASRM | n (%) | I | 12 (100) | - | - |  |
|  |  | II | 0 (0) | - |  |  |
|  |  | III | 0 (0) | - |  |  |
|  |  | IV | 0 (0) | - |  |  |
| SECRETORY GROUP |  |  |  |  |  |  |
| Characteristic | Unit | Detail | Patients with PE | Patients with PE and OE | Controls | p-value |
| Number of patients | n | - | 8 | 8 | 8 |  |
| Age (mean ± SD) | years | - | 29.13 ± 4.02 | 31.5 ± 2.62 | 30.75 ± 2.12 | 0.297 |
| BMI (mean ± SD) | kg/m<sup>2</sup> | - | 22.91 ± 2.07 | 21.45 ± 1.87 | 21.91 ± 2.86 | 0.448 |
| Oral contraceptives in the last 3 months | n (%) | Yes | 0 (0) | 0 (0) | 0 (0) | >0.999* |
|  |  | No | 8 (100) | 8 (100) | 8 (100) | >0.999** |
| Hormonal therapy in the last 3 months | n (%) | Yes | 1 (12.5) | 0 (0) | 0 (0) | >0.999* |
|  |  | No | 7 (87.5) | 8 (100) | 8 (100) | >0.999** |
| Medication in the last week | n (%) | Yes | 4 (50) | 3 (38) | 2 (25) | 0.608* |
|  |  | No | 4 (50) | 5 (62) | 6 (75) | 0.999** |
| Smoking status | n (%) | Non-smoker | 5 (62.5) | 5 (62) | 3 (37.5) | 0.246*<br>0.888** |
|  |  | Smoker | 2 (25) | 0 (0) | 1 (12.5) |  |
|  |  | Occasional smoker | 0 (0) | 0 (0) | 2 (25) |  |
|  |  | Former smoker | 1 (12.5) | 3 (38) | 2 (25) |  |
| rASRM | n (%) | I | 6 (75) | 0 (0) | - | - |
|  |  | II | 1 (12.5) | 4 (50) | - |  |
|  |  | III | 1 (12.5) | 0 (0) | - |  |
|  |  | IV | 0 (0) | 4 (50) | - |  |

**Abbreviations:** n, number; SD, standard deviation; BMI – body mass index, PE, peritoneal endometriosis, OE, ovarian endometriosis; rASRM, revised American Society of Reproductive Medicine score; \*PE *versus* controls, \*\*PE and OE *versus* controls

Study included an untargeted transcriptomic approach for biomarkers discovery, combined with machine learning techniques. Total RNA was extracted from patients’ whole blood and subjected to ribosomal RNA removal. Quantified cDNA libraries were then prepared, and whole-genome RNA was sequenced using Illumina platforms. The RNA sequencing data were used for differential gene and transcript expression analysis, as well as pathway enrichment analysis. Additionally, the sequencing data were integrated into a machine learning pipeline for the selection of the most informative genes and transcripts using feature selection techniques, followed by the construction of support vector machine classifiers to predict the endometriosis status of patients. The flowchart of the study design is shown in Figure 2.

**Figure 2.**
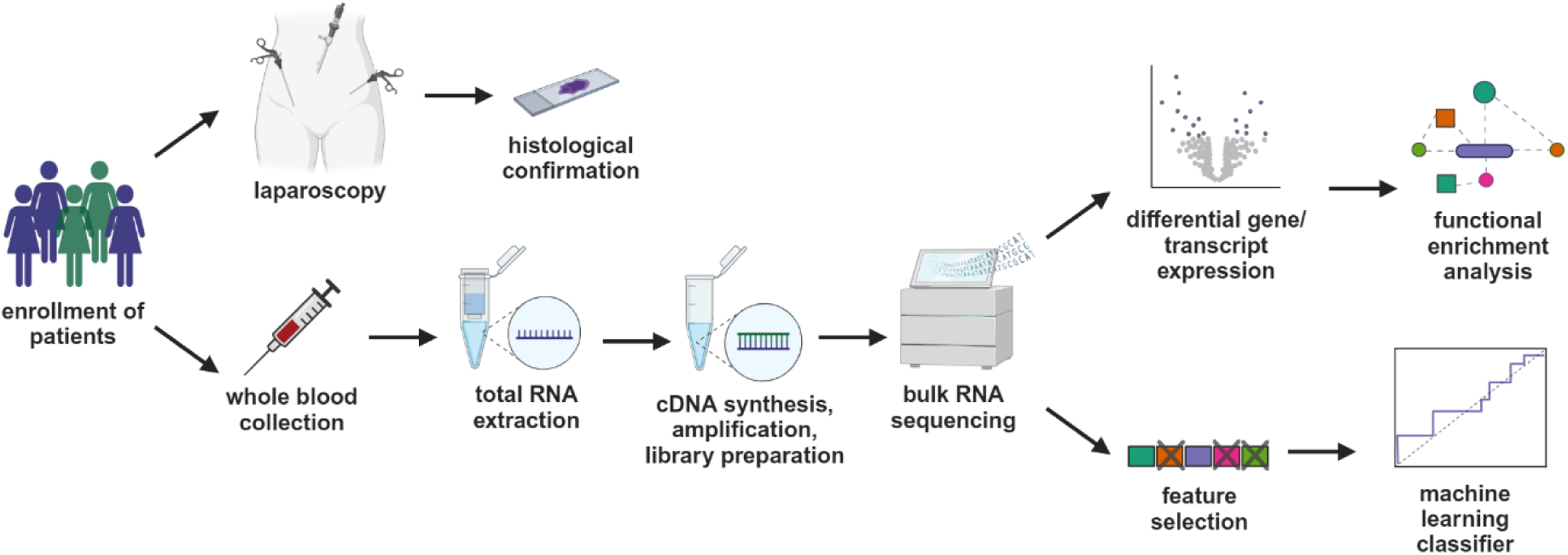
Schematic representation of the study design. Eligible participants were recruited prospectively and underwent laparoscopy, with histological confirmation of endometriotic lesions. Whole blood samples from patients were collected, total RNA was extracted, quantified, and cDNA libraries were prepared and sequenced using Illumina platforms. The RNA sequencing data were used for differential gene and transcript expression analysis, functional enrichment analysis, and were integrated into a machine learning pipeline for feature selection and construction of a classifier to predict the endometriosis status of patients. Created with BioRender.com.

### Sample collection and processing

Sample collection and processing were carried out according to a strict standard operating procedure ^43^. Up to one day to one hour before surgery, 3 ml blood samples were collected into Tempus Blood RNA tube (Applied Biosystems, Waltham, MA, USA) at the Department of Obstetrics and Gynaecology at the UMC Ljubljana, Slovenia and at the Department of Gynaecology and Medical University Centre Vienna, Austria. Immediately after blood collection, the tubes were shaken vigorously for at least 20 seconds. The tubes were stored at 4 ° C for up to 5 days and then transferred to -80°C until further analysis according to manufacturer’s instructions.

### RNA isolation and quality analysis

RNA was isolated according to the manufacturer’s instructions using the Tempus Spin RNA Isolation kit (Applied Biosystems, Waltham, MA, USA). Briefly, stabilised blood was thawed at room temperature and transferred into a 50 ml tube. Then, 3 to 4 ml of 1 x phosphate-buffered saline (PBS) was added to the tube to reach a total volume of 12 ml. The tube was vortexed vigorously for 1.5 minutes and centrifuged at 3000 × g for 30 minutes at 4 °C. After centrifugation, the tube contents were carefully poured off, and the RNA pellet was resuspended in 400 µl of RNA Purification Resuspension Solution. The resuspended RNA was transferred to a purification filter and purified with Wash Solutions 1 and 2. Finally, the RNA was eluted in Nucleic Acid Purification Elution Solution, aliquoted and stored at -80 °C for further analysis. The concentration of isolated RNA was determined using a NanoDrop One (Thermo Fisher Scientific, Waltham, MA, USA), while RNA integrity was assessed with the Agilent Bioanalyzer Instrument using the RNA 600 Nanokit (Agilent Technologies Inc., Santa Clara, CA, USA). RNA samples were subjected to strict quality control before being sent for sequencing. Samples with an RNA Integrity Number (RIN) > 7.5, a concentration of at least 20 ng/µl, and a volume of 20 µl were used for whole genome RNA sequencing (Novogene, Cambridge, UK).

### RNA sequencing

First, ribosomal RNA was removed and rRNA free residues were cleaned using an rRNA removal kit and ethanol precipitation. Subsequently, RNA fragmentation was performed, and first-strand cDNA was synthesised. During second-strand cDNA synthesis, dTTPs were replaced by dUTPs in the reaction buffer. The directional library was prepared after end repair, A-tailing, adapter ligation, size selection, USER enzyme digestion, amplification, and purification. Quantified libraries were pooled and sequenced on Illumina platforms. The samples were sequenced in two separate batches: the first comprising samples from patients in the proliferative group, and the second from those in the secretory group. For alignment and calculation of read counts, we used CLC Genomics Workbench 21.0.4 and 22.0.1 (QIAGEN Aarhus, Denmark) using the Identify and Annotate Differentially Expressed Genes and Pathways 1.19 and RNA-Seq Analysis 2.7 tool with the default settings. We used the reference Homo sapiens GRCh38.104 (Gene, RNA). For calculation of TPM values, we used the Differential Expression for RNA-seq 2.7 and 2.8 tools with the default settings.

### RNA sequencing analysis

We performed differential gene expression (DGE) and differential transcript expression (DTE) analyses based on read counts data using R ^44^ version 4.3.0 with DESeq2 ^45^ version 1.42.1. Prior to analysis, genes/transcripts with a non-zero read count in fewer than six samples were filtered out. The analyses were conducted separately for patients in each of the two menstrual cycle phases. For both phases, we compared patients without endometriosis (controls) to those with endometriosis, grouping patients with PE only and PE and OE together. Additionally, for the secretory phase, we performed a further analysis by dividing patients into three groups: (1) patients without endometriosis, (2) patients with PE only, and (3) patients with both PE and OE. We then compared each group against the other two groups separately. Genes/transcripts were considered differentially expressed if their adjusted p-value was below 0.05. Volcano plots displaying the -log_10_ adjusted p-value against the log_2_ fold change were created using the Matplotlib library. The workflow of the experimental analysis is illustrated in Figure 3.

**Figure 3.**
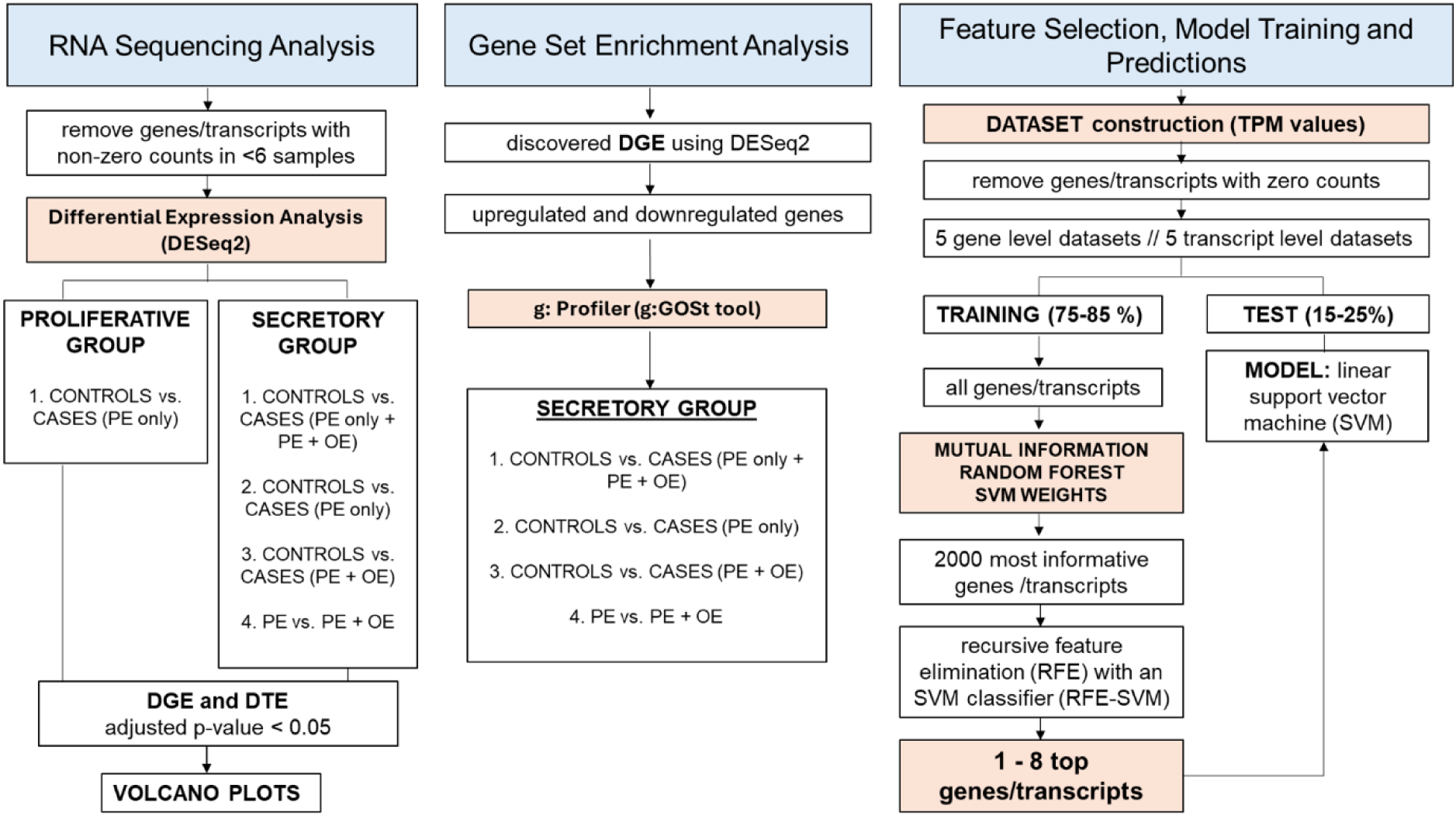
Scheme of experimental workflow. PE – peritoneal endometriosis, OE – ovarian endometriosis, DGE – differential expressed genes, DTE - differentially expressed transcripts, TPM – transcripts per million, SVM – support vector machine.

### Gene set enrichment analysis

Only DEGs identified in patients from the secretory group were included in the gene set enrichment analysis. The DGEs were compared across four groups: controls vs PE only, controls vs PE + OE, controls vs all cases (PE only and PE + OE), and PE only vs PE + OE (Figure 3). All DGEs were divided into two subsets: upregulated and downregulated genes. Within each subset, genes were ranked in ascending order according to their adjusted p-values. These ranked gene lists were then individually inputted into the g:GOSt tool, a component of the g:Profiler web service ^46^, for gene set enrichment analysis. The “ordered query” option was selected, while other settings remained at their default values. The analysis was performed using g: Profiler version e113_eg59_p19_f6a03c19, with the database last updated on Mon Jun 16 2025 ^46^.

### Principal component analysis

Genes/transcripts with zero entries across all samples were filtered out. Transcript per million (TPM) values were centred based on genes/transcripts and log-transformed before conducting principal component analysis (PCA). PCA was separately conducted for each menstrual cycle phase as well as for the combined data of all patients across both phases. This analysis was performed using the scikit-learn library for dimensionality reduction and with the Matplotlib library for visualization.

### Feature Selection, Model Training, and Predictions Datasets

For this part, transcript per million (TPM) values were used. We constructed ten datasets: five at the gene level and five at the transcript level. The gene-level datasets included: 1) all participants in the proliferative phase, 2) all participants in the secretory phase, 3) all participants (both proliferative and secretory phases), 4) participants in the secretory phase restricted to controls and cases with PE only, and 5) participants in the proliferative phase restricted to a controls and cases with PE only (Figure 3). The transcript-level datasets were constructed using the same participant groupings as the gene-level datasets.

First, the genes/transcripts with zero entries across all samples were removed. Second, each dataset was divided into training and test sets. In datasets containing participants from only one menstrual cycle phase, 25% of participants were allocated to the test set. In contrast, for datasets with participants from both phases, 15% of participants were assigned to the test set. The test sets were formed by randomly sampling an equal number of participants from each group to maintain the original proportions between groups in the dataset. By ‘group’, we refer to patients in the same menstrual phase and with the same endometriosis status (absence of endometriosis, PE, or PE + OE).

### Feature selection on the training set

Consistent with the machine learning terminology, we refer here to each gene/transcript as a feature. Feature standardization was performed to normalize the data, transforming it to have a mean of zero and a standard deviation of one. Then, the importance of each feature was assessed by applying three different techniques (separately to each of the ten training datasets): mutual information, random forest importance and support vector machine (SVM) weights. All three were implemented using the scikit-learn library. Mutual Information assessed the mutual dependence between each feature and the endometriosis status. For random forest importance, feature importance was evaluated based on how effectively each feature improved the predictive accuracy of a trained random forest classifier. For SVM weights, a linear SVM model was trained on all features and then, the weights of the features, indicating their influence on the decision boundary, were extracted. From the results of each feature selection technique, a list of the 2000 most important genes/transcripts was prepared. Using this shortlist, recursive feature elimination (RFE) with an SVM classifier (RFE-SVM) ^47^ was then performed to further refine the selection of relevant features. Initially, an SVM classifier was trained on data from genes/transcripts from the list, and its performance was assessed through leave-one-out cross-validation on the training dataset, calculating the area under the curve (AUC) of the receiver-operator characteristic (ROC) curve. Subsequently, genes/transcripts were sorted based on their contribution to distinguishing patients with endometriosis from those without it, and the least informative gene/transcript was removed from the list. This iterative process continued until the AUC started decreasing, i.e. at the end of this process, the minimal set of genes/transcripts that achieved an AUC of 1.0 on the training set (indicating a perfect classifier capable of accurately categorizing patients with or without endometriosis across all thresholds) was retrieved.

### Predictions on the test set

To evaluate the performance of the models on unseen data, each set of genes/transcripts obtained from RFE was used for training the SVM model. The training set was filtered to exclusively include genes/transcripts from the retrieved set, and likewise, the test set was filtered accordingly. Subsequently, sensitivity, specificity, and the area under the curve (AUC) of the ROC curve were computed. From the model’s decision function, ROC curves were calculated with the library pROC in R and visualized with the Matplotlib library in Python.

### SVM model hyperparameter tunning

For the linear SVM, the only hyperparameter needing selection is C. We opted for tuning C using the training set comprising participants in the secretory phase, filtered to retain solely differentially expressed transcripts. This decision stemmed from the dataset’s apparent linear separability observed on the PCA plot. We evaluated C across a range of values: 0.001, 0.01, 0.1, 1, 10, and 100, calculating the ROC AUC via leave-one-out cross-validation on the same dataset. From this analysis C value of 0.1 was selected for further utilization, being the second smallest value associated with an AUC of 1.0.

### Statistical analysis of patient’s clinical data

Patients’ clinical data were analysed as follows. The normality of distribution was assessed using the Shapiro-Wilk test, and the outliers were identified and excluded using the ROUT method (Q = 1, FDR < 1 %). For continuous variables, the unpaired t-test or Mann-Whitney test was used. Categorical clinical variables were compared using Fisher’s exact test, the Chi-squared test, or Chi-squared test for trend. Statistical analysis was performed with GraphPad Prism 9.3 (GraphPad Software, San Diego, CA, USA), with the significance level set at p < 0.05.

## RESULTS

### Clinical characteristics of patients

The study included two groups: the proliferative group and the secretory group. In the proliferative group, 12 patients with PE and 12 control subjects were included. The secretory group included 8 patients with PE, 8 patients with both PE and OE, and 8 controls (Figure 1).

In both the proliferative and secretory group, there were no significant differences between cases and controls regarding age, body mass index (BMI), smoking status or use of oral contraceptives or hormonal therapy in the last 3 months before surgery. A statistically significant difference was observed in medication use prior to surgery between controls and cases only in the proliferative group (p=0.014). All endometriosis patients in the proliferative group were classified as rASRM stage I. In the secretory group, 75 % of patients with PE were classified as stage I, 12.5% as stage II, and 12.5% as stage III, while none were in stage IV. Among patients with PE and OE group, 50 % were classified as stage II and 50 % as stage IV. Clinical characteristics of patients included in the study is shown in Table 1.

### PCA analysis based on all genes and transcripts clustered patients according to their menstrual phase

PCA was carried out for each menstrual cycle phase individually and for the combined data from all patients in both the proliferative and secretory phases. We visualized the first six principal components against each other, with data points colored according to various factors including endometriosis status, menstrual phase, and various metadata. The metadata encompassed potential technical sources of variation such as recruitment location (Slovenia or Austria), RNA isolation date, time between hospitalization date, as well as clinical and lifestyle information about patients. This patient clinical and lifestyle data comprised the rASRM score, age, age at menarche, maternal and paternal ethnic origins, smoking status, sport/recreation, reports about pelvic, abdominal or back pain, menstrual pain frequency, menstrual pain intensity, pain during sexual intercourse (in general and in the last 3 months), score of pain during sexual intercourses, pain during urination/defecation (in the last 3 months), nausea and vomiting, regularity of menstrual cycle, partus, miscarriage.

When performing PCA on all genes/transcripts, clustering of samples was observed when genes/transcripts from both phases were clustered by menstrual phase (Supplementary Figures 1 and 2). No clustering was observed based on endometriosis status or other metadata (Supplementary Figure 1 - 3).

### In the secretory group, most of the differentially expressed genes and transcripts were identified between controls and patients with peritoneal endometriosis

DGE and DTE analyses were performed separately for each menstrual cycle phase. For the proliferative phase, we compared controls to PE only cases. For the secretory phase, we compared four groups: controls, PE only, PE + OE, and all cases. No DGEs were identified among participants in the proliferative group (Supplementary Figure 4, left). In the secretory group, 48 DGEs were identified between all controls and all cases (Figure 4, left), 1035 DGEs between controls and cases with PE only (Figure 4, middle), 3 DGEs between controls and cases with PE + OE (Figure 4, right), and 16 DGEs between cases with PE only and cases with both PE and PE + OE (Supplementary Figure 4, right).

**Figure 4.**
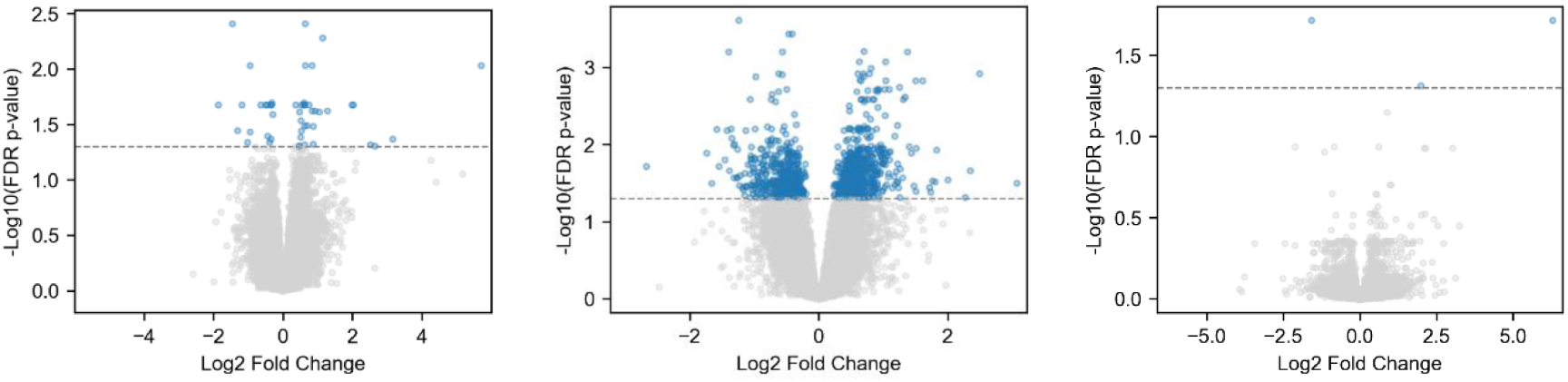
Volcano plots of differentially expressed genes across participants in the secretory group. Volcano plots display differentially expressed genes (DGEs) for the following group comparisons: all controls versus all cases (left), controls versus cases with PE only (middle), and controls versus cases with both PE and OE (right). PE – peritoneal endometriosis, OE – ovarian endometriosis. Differentially expressed genes are located above the dashed vertical line and are colored in blue.

In the proliferative group, only two differentially expressed transcripts were identified (Supplementary Figure 5). In the secretory group, 110 DTEs were identified between all cases and controls (Figure 5, left), 922 DTEs between controls and cases with PE only (Figure 5, middle), 29 DTEs between controls and cases with both PE and OE (Figure 5, right), and 53 DTEs between the cases with PE only and cases with PE and OE (Supplementary Figure 5).

**Figure 5.**
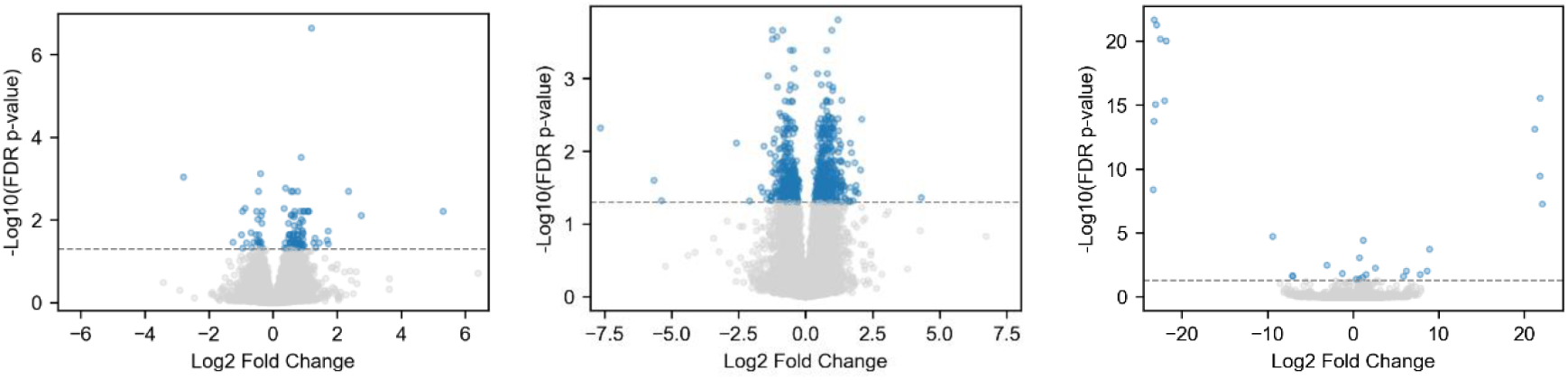
Volcano plots of differentially expressed transcripts across participants in the secretory group. Volcano plot display differentially expressed transcripts for the following group comparisons: all controls versus all cases (left), controls versus cases with PE only (middle), and controls versus cases with both PE and OE (right). PE – peritoneal endometriosis; OE – ovarian endometriosis. Differentially expressed transcripts are located above the dashed vertical line and colored in blue.

### Gene set enrichment analysis highlights immune and inflammatory pathways in peritoneal endometriosis

Gene set enrichment analysis was performed only for the secretory group, using the identified upregulated and downregulated DGEs for comparisons between controls and defined case groups (all cases, PE only, PE + OE).

The highest number of DGEs was identified between controls and cases with PE only. Functional enrichment analysis of the upregulated genes in this group revealed significant enrichment across Gene Ontology (GO) biological processes, Reactome, and KEGG pathways. The top enriched terms ranked by number of DGEs, were related to immune and inflammatory processes such as immune system, innate immune system, neutrophil degranulation, cytokine receptor signalling, and regulation of RAGE receptor binding. Additionally, pathways involved in intracellular signalling, cellular response to stimulus or stress, and vesicle trafficking were also prominent (Figure 6A, B). Downregulated DGEs for the comparison of the same groups were enriched in pathways related to T cell activation, a key component of the cell-based immune response (Figure 6C). Identified enriched pathways for the up and downregulated genes between the following groups: controls, PE only, PE+ OE, all cases (PE + PE +OE), are listed in Supplementary Tables 1 and 2.

**Figure 6.**
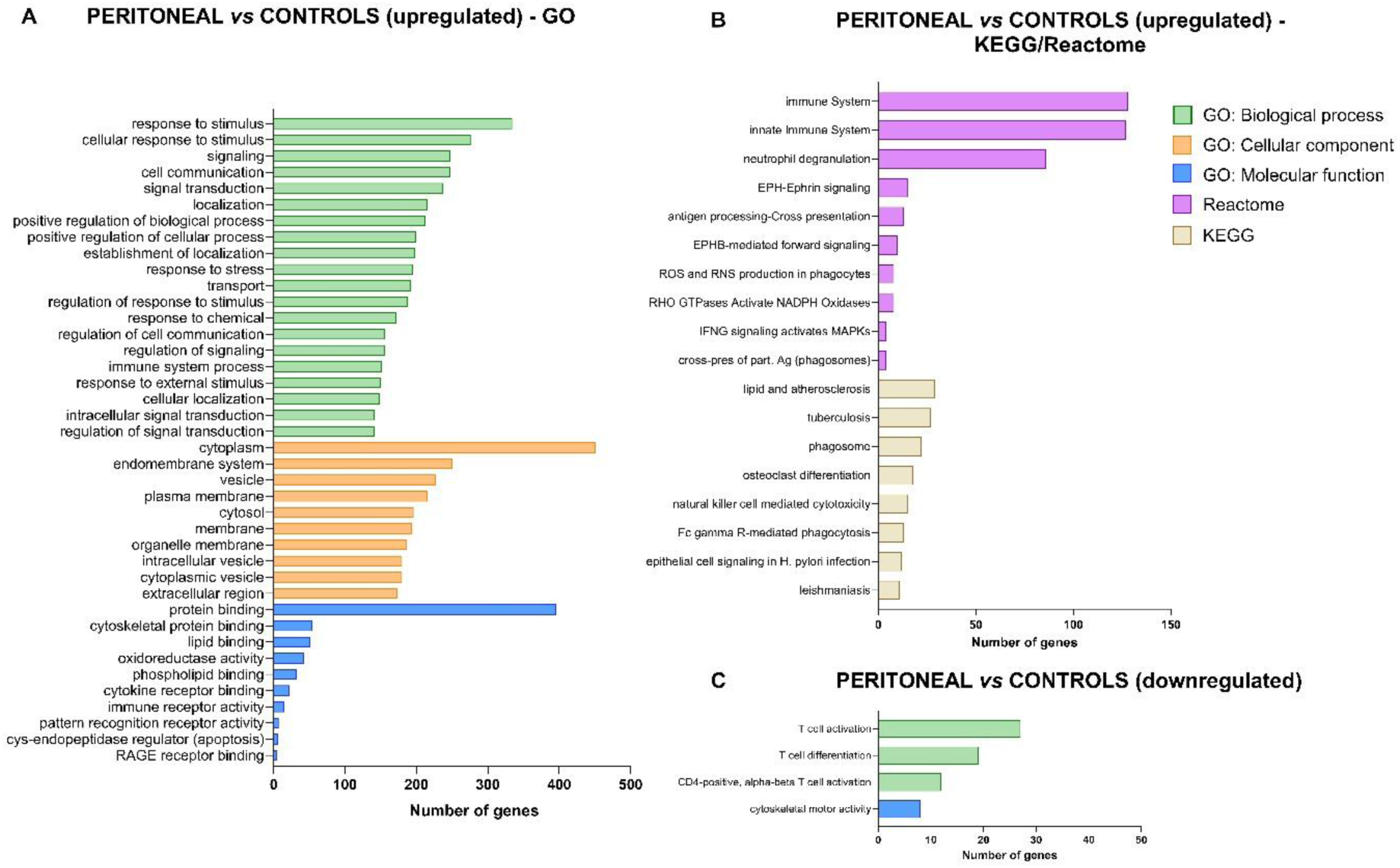
Top enriched pathways in secretory group participants with peritoneal endometriosis. Top enriched pathways are ranked by the number of differentially expressed genes (DGEs) contributing to each term, identified between peritoneal endometriosis patients and controls in the secretory group. Pathway enrichment was performed using g:Profiler. The Y-axis lists the top 10–20 enriched terms, and the X-axis indicates the number of DGEs associated with each pathway. Pathways are ranked by gene count, and only statistically significant terms (adjusted p-value < 0.05) are included. Enrichment analysis is shown for A), B) upregulated and C) downregulated genes.

Among the DGEs identified between all controls and cases, three upregulated genes (*CD93, CXCL8, NINJ1*) were found to be enriched in pathways like angiogenesis, a process known to contribute to the development and progression of endometriosis (Supplementary Table 11). Additionally, two to three upregulated genes (*NINJ1, CD14, PTAFR*) were associated with lipopolysaccharide (LPS) binding and LPS immune receptor activity, which are linked to innate immune responses (Supplementary Table 2). One of the three DGEs identified between controls and cases with both PE and OE endometriosis was *CDO1*, which is associated with taurine and hypotaurine metabolism (Supplementary Table 2). Among the downregulated DGEs identified between cases with PE only and those with both PE and OE, pathway enrichment analysis revealed associations with neutrophil degranulation (*CAMP, CRISP3, ARG1, KRT1*), defence response (*CAMP, CRISP3, ARG1, KRT1, OASL*), and innate immune response (*CAMP, CRISP3, ARG1, KRT1, OASL*) (Supplementary Table 1).

Venn diagram analysis revealed one downregulated gene (*B3GAT1*) that was common in three group comparisons: all controls vs. all cases, all controls vs. PE cases, and all controls vs. PE+ OE cases (Supplementary Figure 6, left). Among upregulated genes, 26 were shared between all controls vs. all cases and all controls vs. PE cases, while one downregulated gene (*CDO1*) overlapped between all controls vs. all cases and all controls vs. PE + OE endometriosis cases (Supplementary Figure 6, right). These overlaps highlight subsets of genes that may be consistently dysregulated across clinical presentations of PE and patients with both PE and OE, with potential relevance to common disease mechanisms and biomarker development.

### Feature selection and model predictions analysis identified a set of six DTEs with the highest discriminatory performance in distinguishing cases from controls in both menstrual phases

Table 2 and Table 3 show the sets of genes/transcripts for which the SVM models achieved the highest ROC AUC values on the test set within each group. ROC curves for the overall best-performing models are shown in Figures 7 and 8, while ROC curves for the remaining top models are provided in Supplementary Figures 7 to 14. In Table 2, for all participants in the proliferative group, a ROC AUC of 0.67, sensitivity of 1.0, and specificity of 0.67 were achieved with the selected 3 genes. For all participants in the secretory group, predictions based on the identified 3 genes resulted in a ROC AUC of 0.88, sensitivity of 0.75, and specificity of 1.0. In the secretory group, for controls and cases with PE only, performance with the selected single gene was slightly worse: ROC AUC of 0.75, sensitivity of 0.5, and specificity of 0.5. When all participants from the proliferative and secretory group were analysed together, the identified set of 8 genes resulted in a ROC AUC of 0.75, sensitivity of 1.0, and specificity of 0.67. For the comparison between all controls and all cases with PE only, the identified set of 7 genes resulted in a lower ROC AUC (0.67) and sensitivity (0.33), but a higher specificity of 1.0.

**Figure 7.**
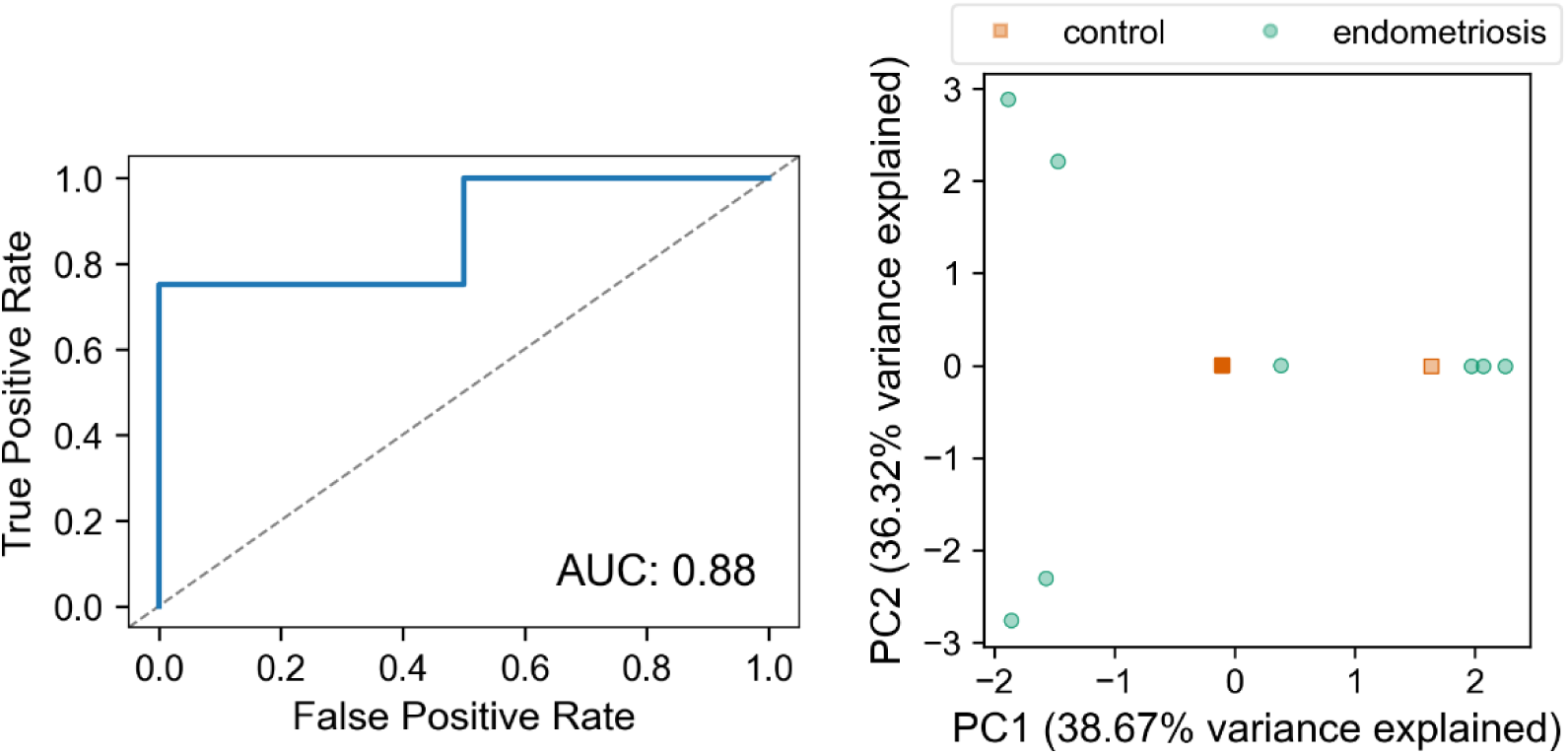
Classification performance and PCA visualization based on selected genes in the secretory group. Left: Receiver operating characteristic (ROC) curve showing the predictive performance of the SVM model for all participants in the secretory group. The model was trained using the gene set identified through the feature-selection procedure initiated with mutual information. Right: Principal component analysis (PCA) plot generated using the same selected gene set for all participants in the secretory group (right).

**Figure 8.**
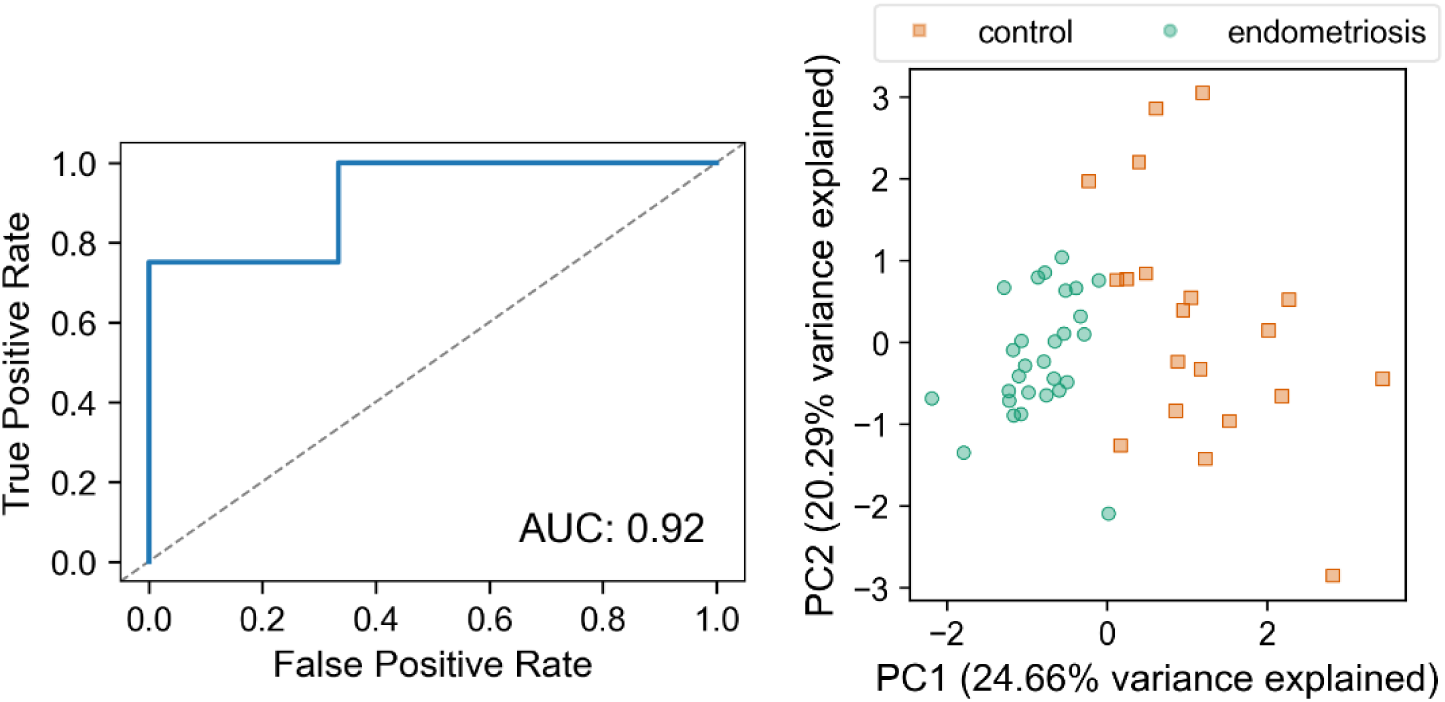
Classification performance and PCA visualization based on selected transcripts for all the participants from both proliferative and secretory group. Left: Receiver operating characteristic (ROC) curve showing the predictive performance of the SVM model for all participants. The model was trained using the set of transcripts identified through the feature-selection procedure initiated with random forest importance. Right: Principal component analysis (PCA) plot generated using the same selected transcript set for all participants.

**Table 2.** Sets of genes identified by the feature selection pipelines for which the SVM models achieved the highest ROC AUC values on the test set within each group.

| Group of participants | Feature selection method prior RFE-SVM | Models based on genes | Performance on test set |
| --- | --- | --- | --- |
| all participants<br>(PROLIFERATIVE GROUP) | mutual information | Gene model 1 (3 genes) | AUC: 0.67<br>Sensitivity: 1.0<br>Specificity: 0.67 |
| **all participants**<br>**(SECRETORY GROUP)** | **mutual information** | **Gene model 2 (3 genes)** | **AUC: 0.88**<br>**Sensitivity: 0.75**<br>**Specificity: 1.0** |
| all controls and cases with<br>peritoneal endometriosis only<br>(SECRETORY GROUP) | random forest<br>importance | Gene model 3 (1 gene) | AUC: 0.75<br>Sensitivity: 0.5<br>Specificity: 0.5 |
| all participants<br>(PROLIFERATIVE AND<br>SECRETORY GROUP) | mutual information | Gene model 4 (8 genes) | AUC: 0.75<br>Sensitivity: 0.75<br>Specificity: 0.67 |
| all controls and cases with<br>peritoneal endometriosis only<br>(PROLIFERATIVE AND<br>SECRETORY GROUP) | SVM weights | Gene model 5 (7 genes) | AUC: 0.67<br>Sensitivity: 0.33<br>Specificity: 0.67 |

\*Model achieving the highest ROC AUC is highlighted bold

**Table 3.** Sets of transcripts identified by the feature selection pipelines for which the SVM models achieved the highest ROC AUC values on the test set within each group.

| Group of participants | Feature selection method prior RFE-SVM | Models based on transcripts | Performance on test set |
| --- | --- | --- | --- |
| all participants (PROLIFERATIVE GROUP) | mutual information/<br>SVM weights | Transcript model 1 (1 transcript) | AUC: 0.78<br>Sensitivity: 0.67<br>Specificity: 0.67 |
| all participants (SECRETORY GROUP) | random forest importance | Transcript model 2 (2 transcripts) | AUC: 0.75<br>Sensitivity: 0.75<br>Specificity: 0.5 |
| all controls and cases with peritoneal endometriosis only (SECRETORY GROUP) | SVM weights | Transcript model 3 (1 transcript) | AUC: 1.0<br>Sensitivity: 1.0<br>Specificity: 0.5 |
| **all participants (PROLIFERATIVE AND SECRETORY GROUP)** | **random forest importance** | **Transcript model 4 (6 transcripts)** | **AUC: 0.92**<br>**Sensitivity: 0.75**<br>**Specificity: 1.0** |
| all controls and cases with peritoneal endometriosis only (PROLIFERATIVE AND SECRETORY GROUP) | mutual information | Transcript model 5 (3 transcripts) | AUC: 0.89<br>Sensitivity: 0.67<br>Specificity: 1.0 |

\*Model achieving the highest ROC AUC is highlighted bold

In Table 3, it is shown that for all participants in the proliferative group, the single selected transcript yielded a ROC AUC of 0.78, sensitivity of 0.67, and specificity of 0.67. For all participants in the secretory group, the two identified transcripts achieved a ROC AUC of 0.75, sensitivity of 0.75, and specificity of 0.5. For the comparison between controls and cases with PE only, the performance with the selected single transcript was very good, with a ROC AUC of 1.0, sensitivity of 1.0, and specificity of 0.5. For all participants, from both the proliferative and secretory group, the identified set of 6 transcripts, resulted in a ROC AUC of 0.92, sensitivity of 0.75, and specificity of 1.0. For the comparison between all controls and all cases with PE only, the transcript set consisting of 3 transcripts resulted in a ROC AUC of 0.89, sensitivity of 0.67, and specificity of 1.0.

When performing PCA on the set of genes/transcript with the highest AUC of the ROC curve (Table 2 and 3) on the test set of the SVM models build on sets and transcripts identified by the feature selection procedure, clustering of patients based on endometriosis status was observed (Figures 7 and 8 and Supplementary Figures 7 to 14).

## DISCUSSION

This is the first study to use the whole-blood transcriptomics and machine learning methods to identify novel biomarker candidates specific to PE. The analysis identified a set of six DTEs with the highest discriminatory performance between controls and cases in the test dataset (ROC AUC= 0.92, sensitivity = 75%, specificity = 100%). Similarly, a three gene panel achieved high performance in the secretory group (AUC = 0.88, sensitivity = 75%, specificity = 100%). These findings suggest that, after validation, blood-based transcriptomic markers may offer promising non-invasive tools for the detection of PE.

To date, most transcriptomics studies using machine learning approaches have focused on analysing gene expression data derived from ectopic, eutopic, or healthy endometrial tissue samples, obtained either from the Gene Expression Omnibus (GEO) database or from newly collected samples subjected to microarray analysis ^48–51^. In contrast, fewer studies have explored non-invasive approaches for biomarker discovery in endometriosis. These studies have examined levels of miRNA in serum, plasma, and saliva ^52–54^, lncRNAs in plasma-derived extracellular vesicles ^55^, mRNA expression in menstrual fluid ^56^ or lncRNAs in the serum of endometriosis patients ^57^. Notably, most studies that searched for non-invasive biomarkers for endometriosis have focused on miRNAs; however, these investigations have reported inconsistent results and limited overlap among the identified candidate miRNA biomarkers ^29,58^. One such study developed a salivary signature comprising 109 miRNAs, developed using miRNA sequencing and a machine learning random forest model, and proposed it as a potential diagnostic tool for endometriosis (commonly referred to as Endotest) ^54,59^. Although this signature was recently validated in a multicentre study^60^, it has not yet received approval from national health technology assessment bodies for routine clinical implementation, as further independent, real-world validation outside controlled research settings is still required In another study, Su et al. ^64^ performed biomarker discovery by analysing publicly available GEO datasets and using machine learning algorithms to develop an integrative model for predicting endometriosis, resulting in a nine-gene diagnostic panel. This model was subsequently validated on whole blood samples from endometriosis patients (n=29) and controls (n= 30). However, more than 65% of patients in the validation cohort had rAFS stage III–IV disease, and two of the nine genes showed suboptimal performance. Further validation in a larger, multicentre cohort is therefore required.

In our study, whole blood samples from patients were used to identify candidate biomarkers for endometriosis through a non-targeted transcriptomics approach. Previous studies have primarily employed targeted strategies, such as measuring the levels of specific mRNA molecules in peripheral blood samples ^36,65^. When RNA sequencing techniques were applied, they were mostly performed on separated blood components, such as serum and plasma ^53,66^ or on isolated peripheral blood mononuclear cells ^67^.

As shown in Tables 2 and 3, different feature selection techniques applied prior to RFE-SVM produced distinct sets of genes and transcripts that yielded the best SVM classifier performance across groups on the test dataset. Interestingly, overlap between feature selection techniques was observed only in the transcript-based analysis of all participants in the proliferative group, where mutual information and SVM weights produced an identical final transcript set following RFE-SVM. Furthermore, there was minimal overlap among the identified gene/transcript sets across different groups, with only two genes being selected more than once, and no transcripts recurring between analyses. Neither of these two genes has previously been linked to endometriosis.

In our study, two SVM models based on DGEs and three models based on DTEs demonstrated strong diagnostic potential, achieving an AUC of up to 1.0, and/or meeting the criteria for rule-in or rule-out tests (sensitivity ≥ 95% for rule-out, specificity ≥ 95% for rule-in) on the test set (Tables 2 and 3). The model based on three DGEs showed the best performance in discriminating endometriosis patients from controls in the secretory group, reaching an AUC of 0.88, sensitivity of 75 % and specificity of 100 %. Of these, two genes are long noncoding RNAs (lncRNAs), previously not associated with endometriosis, while one is a small nucleolar RNA (snoRNA), found to be increased in colorectal endometriosis compared to the eutopic endometrium of women with endometriosis ^68^.

Encouragingly, one model based on DTEs performed well even on datasets including participants in both menstrual phases, effectively predicting endometriosis status regardless of the menstrual phase at the time of blood withdrawal. Such a menstrual-phase-independent test would be much more practical for clinical use, as it simplifies and standardises sample collection, eliminating the need to perform the test in a specific menstrual cycle phase. This model, also based on a set of six transcripts, showed the highest performance, reaching an AUC of 0.92, sensitivity of 75 % and specificity of 100 %. Among the panel of six identified transcripts, five are protein-coding, while one is a retained intron.

When performing PCA, we did not detect sources of variation in the data originating from technical variation or clinical data, except for the menstrual phase which in our case coincided with the sequencing batch. Therefore, it was challenging to determine conclusively whether the observed differences in PCA between patients in different phases stem from genuine biological differences were instead a result of technical variations between the two sequencing batches (referred to as batch effects ^69^, and to mitigate this effect computationally. Consequently, gene/transcript differential expression analysis was conducted separately for each menstrual phase. Previous studies have shown that both normal endometrial tissue and endometriotic lesions exhibit phase-dependent variations in gene expression ^70,71^. Therefore, it is important to account for menstrual phase as a variable when trying to identify reliable molecular biomarkers of endometriosis. Transcriptomic profiling of whole blood across menstrual phases in our study revealed distinct, phase-specific expression patterns associated with PE. No DGEs and only two DTEs were detected in samples from the proliferative group. The observed difference in medication use prior to surgery between controls and cases in the proliferative group may have influenced transcriptional profiles, and contributed to the limited number of differentially expressed genes and transcripts detected in this group. In contrast, in the secretory group, tens to hundreds of DGEs and DTEs were identified between cases and controls. Specifically, the highest numbers of DGEs (1035) and DTEs (922) were detected between all controls and PE only cases. No overlapping DGEs or DTEs were observed between the two menstrual phases, underscoring the phase-specific expression profiles.

A common approach in biomarker discovery studies relies on selecting differentially expressed genes based on p-values and/or fold changes. However, this method can miss biologically relevant signals, especially when expression changes are subtle or phase-specific or may yield long lists of genes that are challenging and impractical to use for validation or diagnosis. In addition, analyses at the gene level do not account for transcript-specific changes that may be critical in distinguishing groups. To address these limitations, machine learning methods have been increasingly applied, as they handle complex transcriptomic data and identify patterns that conventional methods might overlook. Machine learning also has limitations, one being multiplicity, where two distinct models achieve similar performance in the training dataset but vary greatly on independent/test datasets. This instability, also observed in our study, indicates that model accuracy alone does not guarantee the identification of biologically relevant features. Consequently, multiplicity can lead to inconsistent biomarker sets, emphasising the need for cautious interpretation and experimental validation ^72^.

This study has several strengths. It analysed the whole blood transcriptome, providing a non-invasive and technically robust approach that avoids pre-analytical variability introduced by plasma or serum processing and captures the full range of RNA species, not limited to mRNA or miRNA. The study focuses on the most common form of endometriosis, PE, which is still not detectable with imaging techniques. Strict standard operating procedures for blood collection and RNA isolation were followed, and cases and controls were carefully matched for age, BMI, hormonal therapy, and smoking status to minimise confounding effects. Because endometriosis is an oestrogen-dependent disease influenced by hormonal fluctuations throughout the menstrual cycle, patients were stratified by menstrual phase. This approach enabled the identification of biomarkers that are independent of cycle phase, as well as markers that are specific to phases of the cycle. The limitations of this study include a relatively small sample size, difference in medication intake between cases and controls in the proliferative group, lack of technical replication across sequencing batches, and partial confounding of menstrual phase with sequencing batch. To minimise technical variability, all samples were processed at the same time by the same operator using identical protocols. Additionally, the findings have not yet been validated in a larger, independent cohort or in populations of different ethnicities and work in progress will address this through qPCR validation of the six identified transcripts.

## CONCLUSION

To the best of our knowledge, this is the first study to integrate whole-genome transcriptomics with machine learning techniques using whole blood samples for discovery of candidate biomarkers associated with PE. Our analysis identified a six-transcript panel that performed well in distinsguishing endometriosis patients from controls. However, validation in larger, independent cohorts is necessary to confirm its diagnostic potential. The study revealed distinct gene expression profiles between patients in the proliferative and secretory phases of the menstrual cycle, confirming the influence of hormonal status on transcriptional patterns. The differentially expressed genes identified as upregulated in PE patients compared to controls were associated with angiogenesis and innate immune pathways, supporrting important role of these processes in the pathophysiology of PE.

## Data Availability

All data produced in the present study are available upon reasonable request to the authors.

## AUTHOR CONTRIBUTION

Conceptualization, T.L.R.; investigation, M.P.N., T.R. and A.V., resources, T.L.R. and H.B.F; data curation, H.B.F., R.W, M.P.N., writing—original draft preparation, M.P.N. and A.V., writing—review and editing, T.L.R., T.R. ; visualization, M.P.N., A.V. and T.R.; supervision, T.L.R.; project administration, T.L.R. funding acquisition, T.L.R. All authors have read and agreed to the published version of the manuscript.

## ACKNOWLEDGEMENTS

The authors thank their study participants, who kindly donated their samples and time and the personnel of the Department of Obstetrics and Gynaecology, University Medical Centre Ljubljana, Ljubljana, Slovenia, especially Mrs. Tatjana Lončar.

