## Supplementary Material_Figures for "Transcriptome profiling to identify blood biomarkers for peritoneal endometriosis"

**Supplementary Figures**

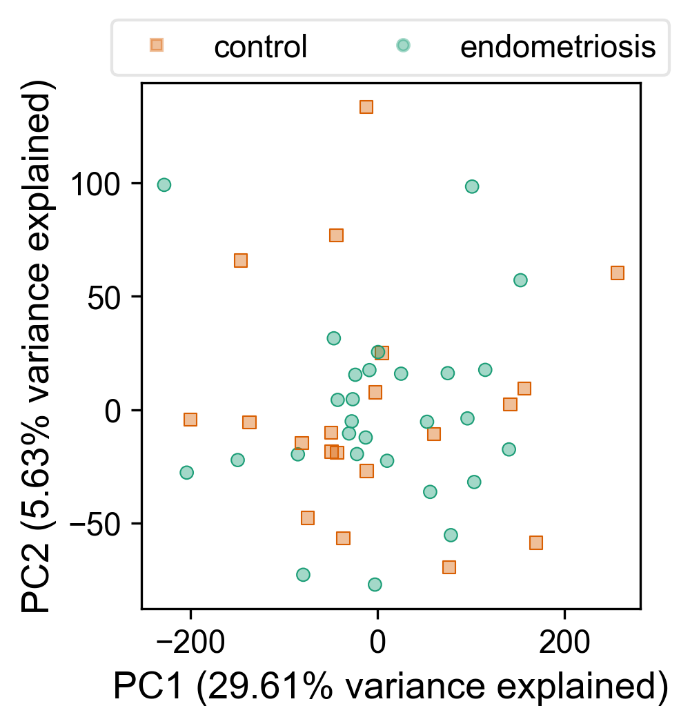

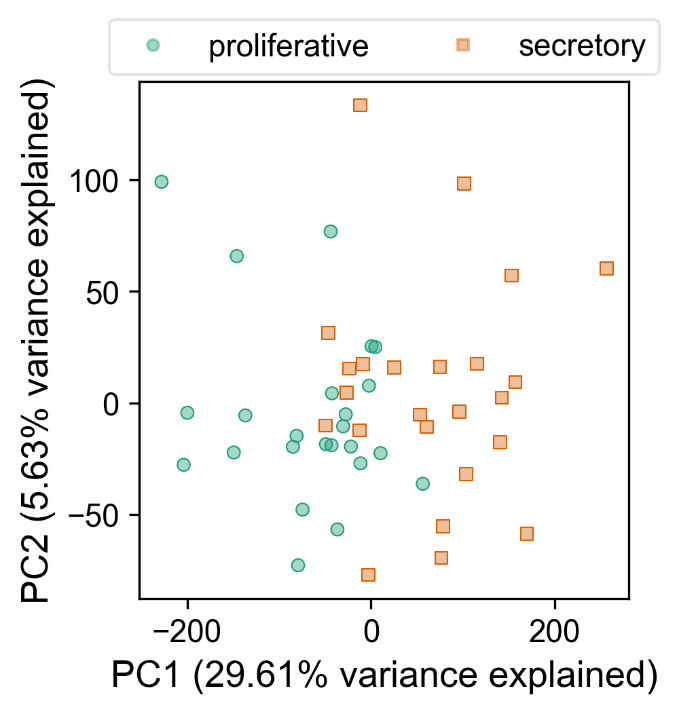

**Supplementary Figure 1.** Principal component analysis of gene expression data. Scatter plots show the first (PC1) and second (PC2) principal components calculated from the expression levels of all genes. Samples are colored according to endometriosis status (left) and menstrual phase (right).

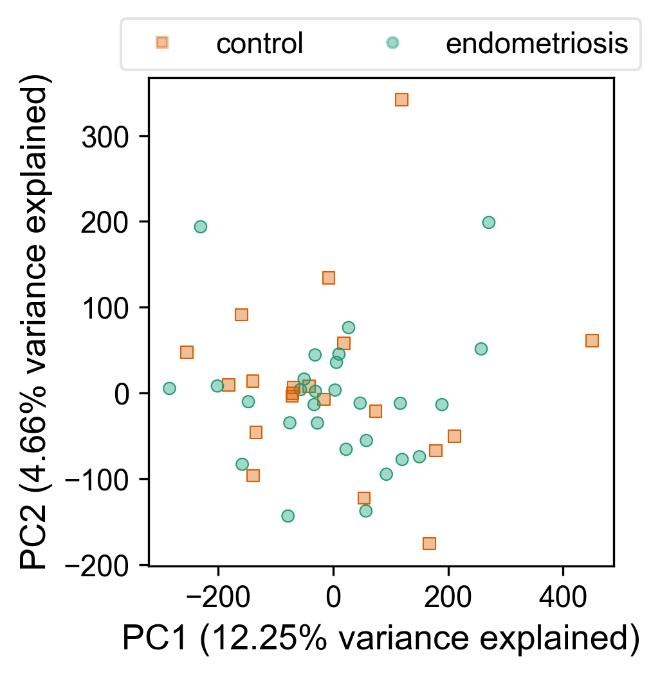

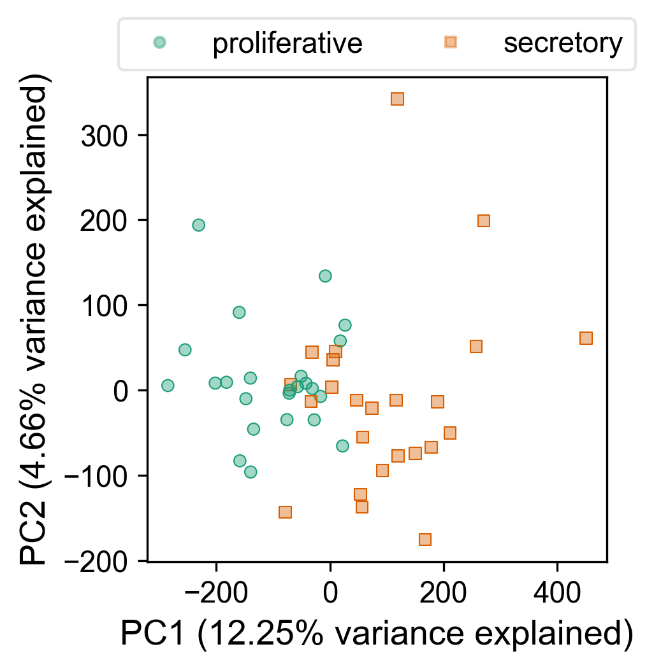

**Supplementary Figure 2.** **Principal component analysis of transcript-level expression data.** Scatter plots display the first (PC1) and second (PC2) principal components calculated from the expression levels of all transcripts. Samples are colored according to endometriosis status (left) and menstrual phase (right).

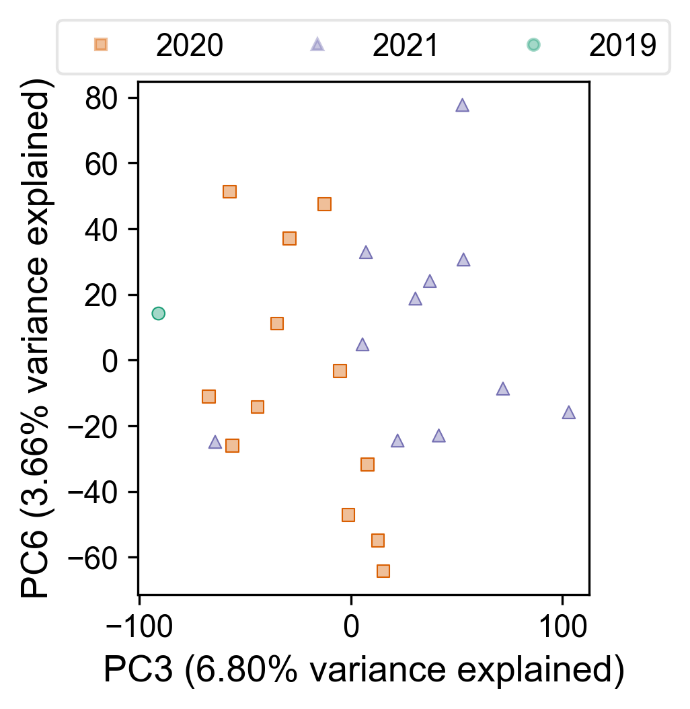

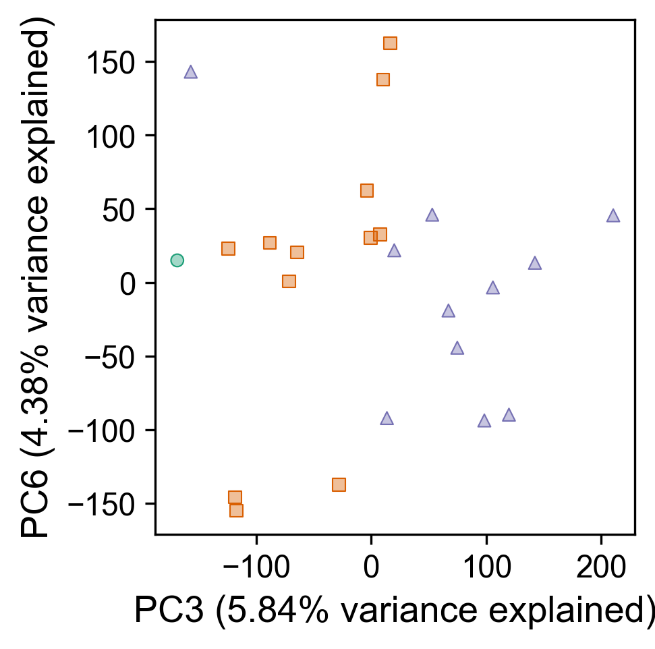

**Supplementary Figure 3.** **Principal component analysis of gene- and transcript-level expression in the secretory phase based on year of hospitalization.** Scatter plots show the third (PC3) and sixth (PC6) principal components calculated from all genes (left) and all transcripts (right) for patients in the secretory phase. Samples are colored according to the year of hospitalization.

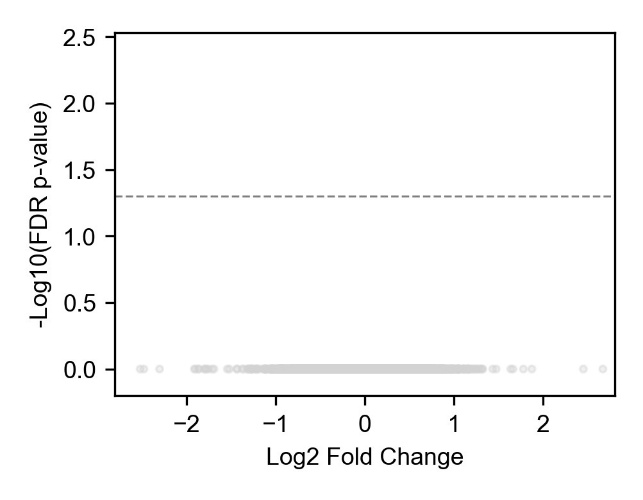

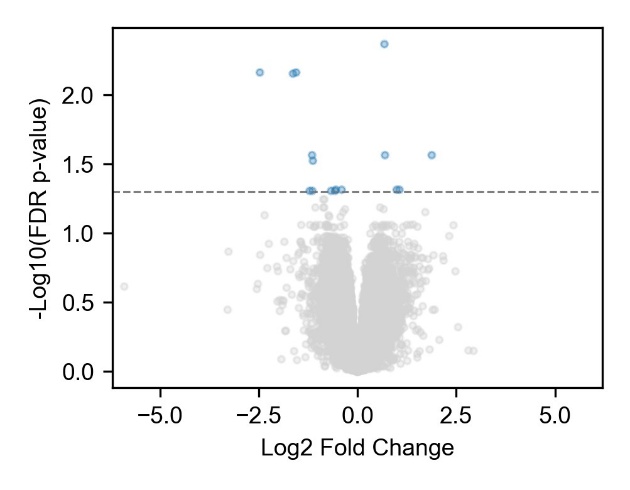

**Supplementary Figure 4.** **Volcano plots of differentially expressed genes in proliferative and secretory groups.** Volcano plots display differentially expressed genes (DGEs) for the following comparisons: controls versus cases in the proliferative group (left), patients with PE only versus PE + OE in the secretory group (right). PE – peritoneal endometriosis, OE – ovarian endometriosis. Genes above the dashed vertical line are considered differentially expressed and are highlighted in blue.

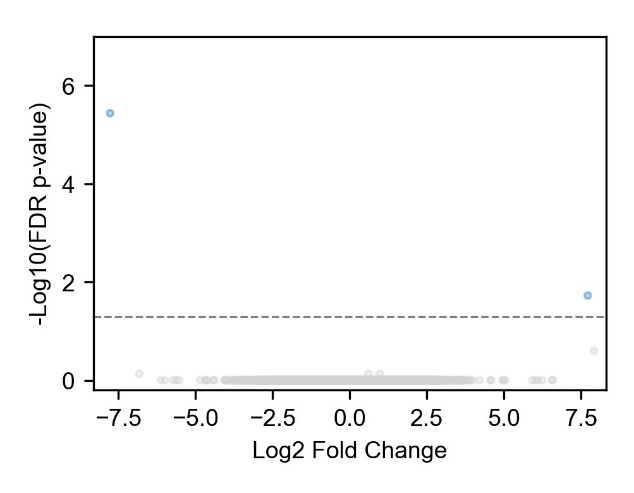

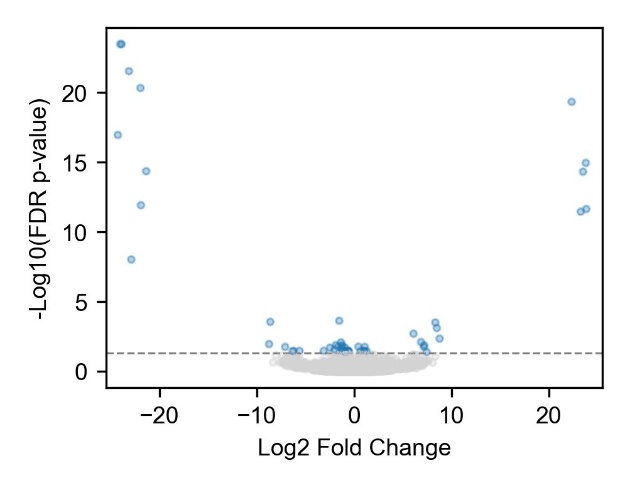

**Supplementary Figure 5.** Volcano plots of differentially expressed transcripts in proliferative and secretory groups. Volcano plots display differentially expressed transcripts (DTEs) for the following comparisons: controls versus cases in the proliferative group (left), patients with PE only versus PE + OE in the secretory group. PE – peritoneal endometriosis, OE – ovarian endometriosis. Transcripts above the dashed vertical line are considered differentially expressed and are highlighted in blue.

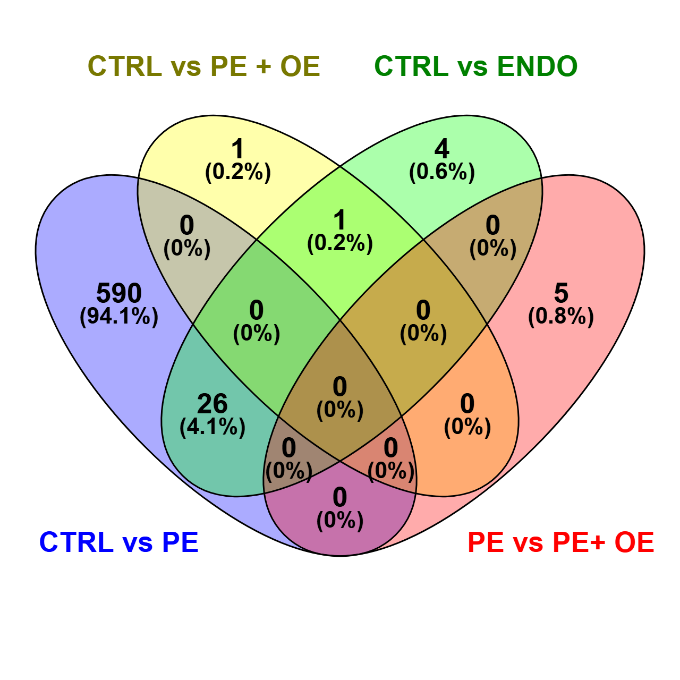

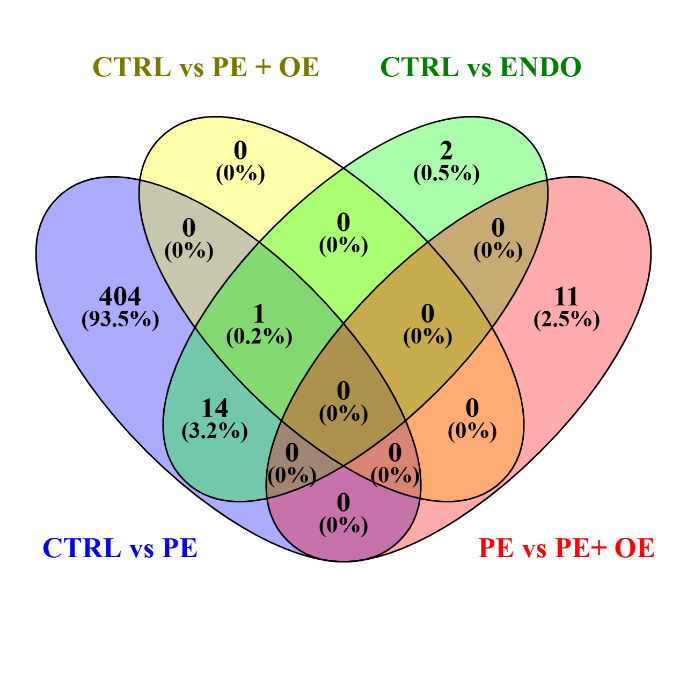

**Supplementary Figure 6.** Venn diagrams showing overlaps of differentially expressed genes (DGEs) across patient subgroups in the secretory phase. The following subgroups were included: controls vs PE only patients, controls vs PE+ OE patients, controls vs all cases, patients with PE only vs patients with PE+ OE. Overlap of upregulated genes (left) and downregulated (right) between each group is shown. Venn diagrams prepared using Venny ^1^. PE – peritoneal endometriosis, OE – ovarian endometriosis.

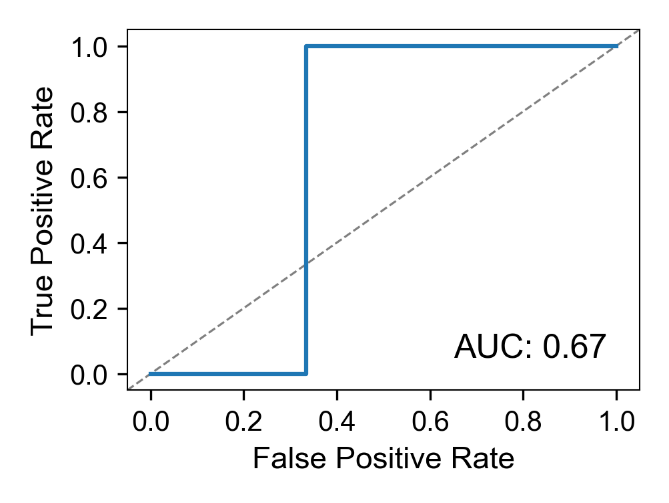

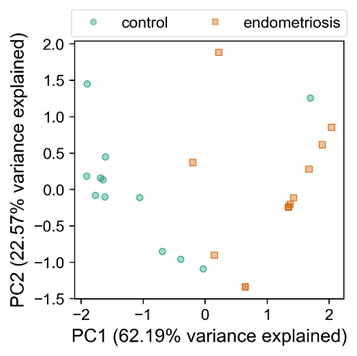

**Supplementary Figure 7.** Classification performance and PCA visualization based on selected genes in proliferative group. Left: Receiver operating characteristic (ROC) curve showing the predictive performance of the SVM model for all participants in the proliferative group. The model was trained using the gene set identified by the feature selection procedure initiated with mutual information. Right: Principal component analysis (PCA) plot constructed using the same selected set of genes for all participants in the proliferative group.

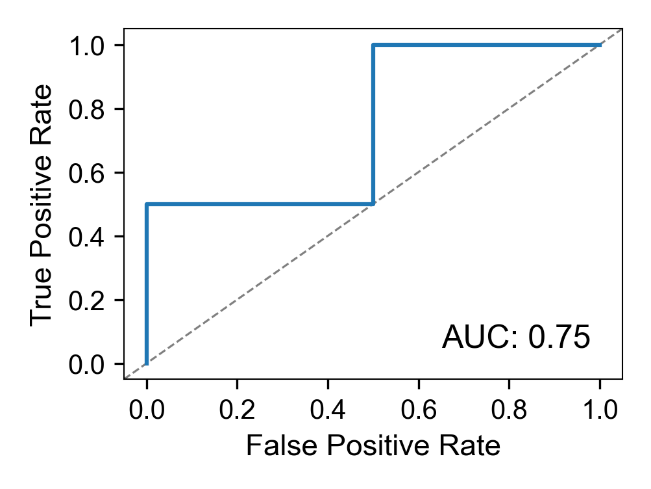

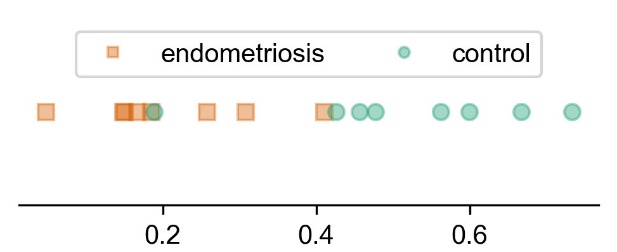

**Supplementary Figure 8**. Classification performance and expression of gene from Model 3 between controls and peritoneal endometriosis patients in secretory group. Left: Receiver operating characteristic (ROC) curve showing the predictive performance of the SVM model distinguishing controls from patients with peritoneal endometriosis only. The model was trained using the gene from Model 3, identified through the feature-selection procedure initiated with random forest importance. Right: Plot of TPM values of gene from Model 3 for the same participants, with points colored according to endometriosis status.

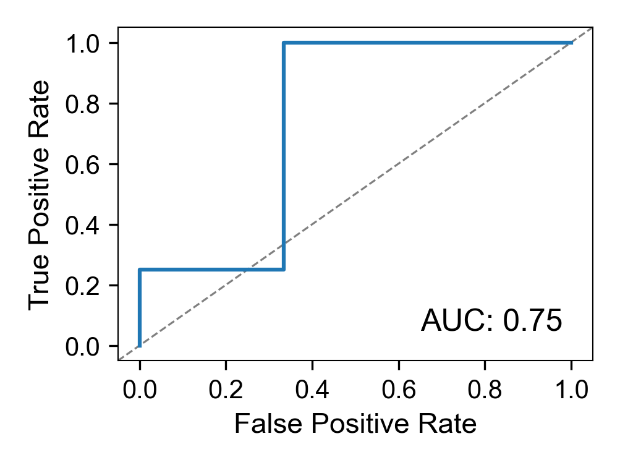

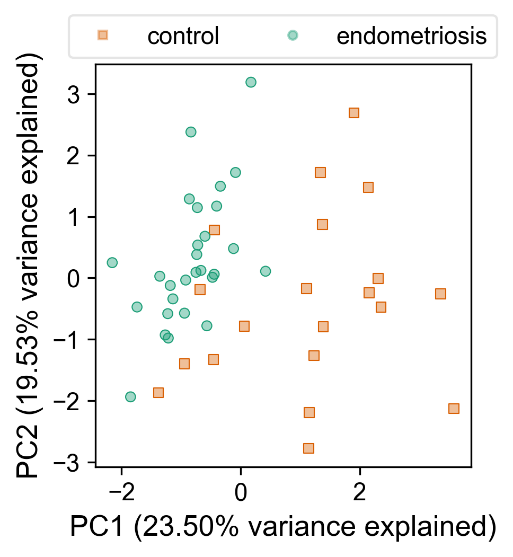

**Supplementary Figure 9.** Classification performance and PCA visualization based on selected genes in participants from both proliferative and secretory group. Left: Receiver operating characteristic (ROC) curve showing the predictive performance of the SVM model for all the participants. The model was trained using the gene set identified through the feature-selection procedure initiated with mutual information. Right: Principal component analysis (PCA) plot generated using the same selected gene set for all participants).

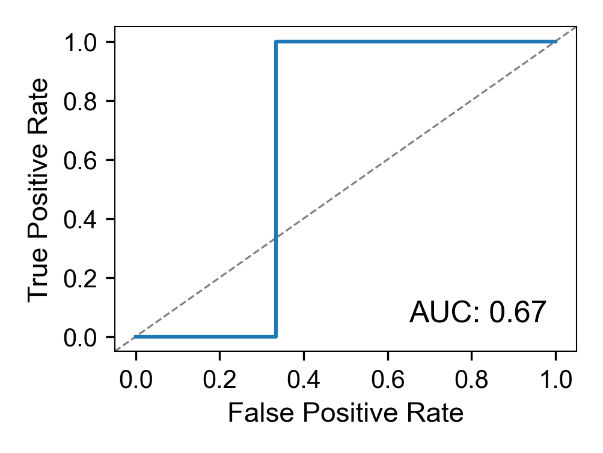

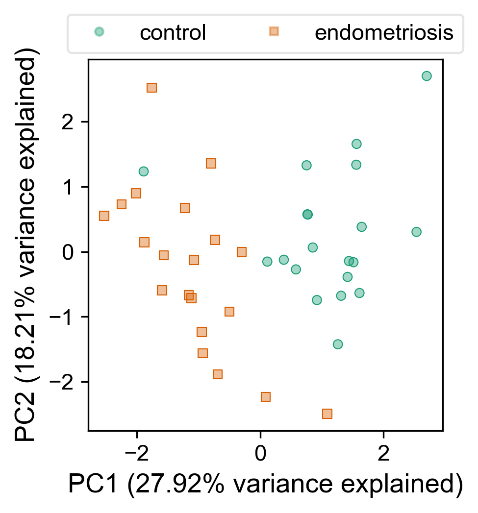

**Supplementary Figure 10**. Classification performance and PCA visualization based on selected genes between all controls and cases with peritoneal endometriosis from both secretory and proliferative group. Left: Receiver operating characteristic (ROC) curve showing the predictive performance of the SVM model distinguishing controls from cases with peritoneal endometriosis only. The model was trained using the gene set identified through the feature-selection procedure initiated with SVM weights. Right: Principal component analysis (PCA) plot generated using the same selected gene set for controls and cases with peritoneal endometriosis only.

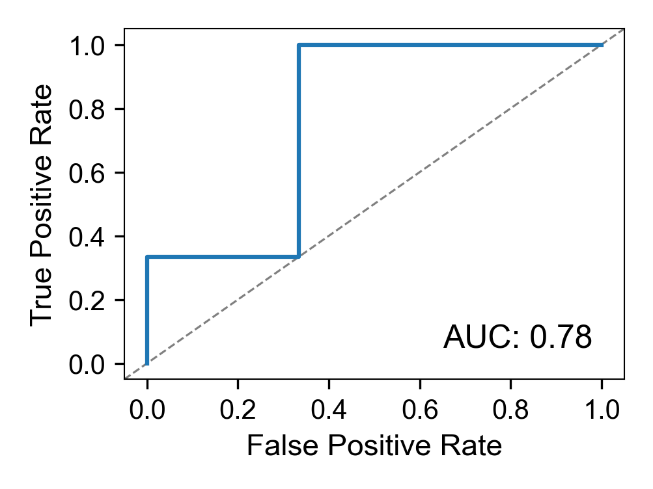

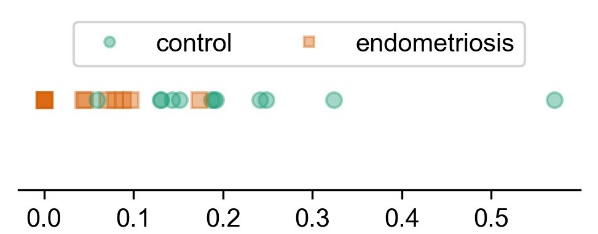

**Supplementary Figure 11.** Classification performance and expression of transcript from transcript model 1 in all participants from the proliferative group. Left: Receiver operating characteristic curve showing the predictive performance of the SVM model for all participants in the proliferative group. The model was trained using the transcript from transcript model 1, identified through the feature-selection procedure initiated with mutual information and SVM weights. Right: Plot of TPM values of transcript from transcript model 1 for the same participants, with points colored according to endometriosis status.

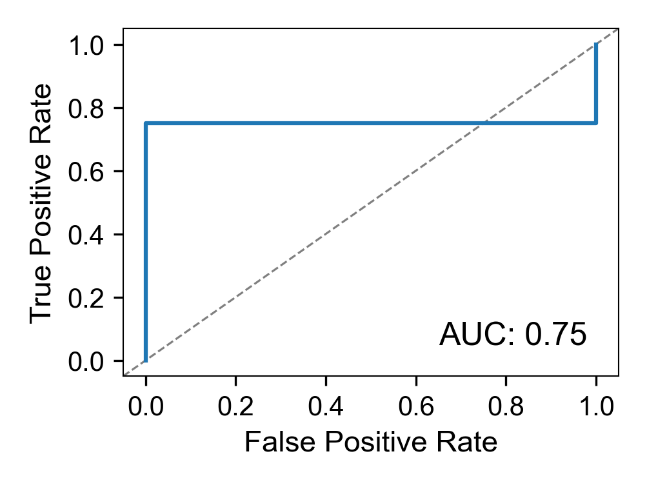

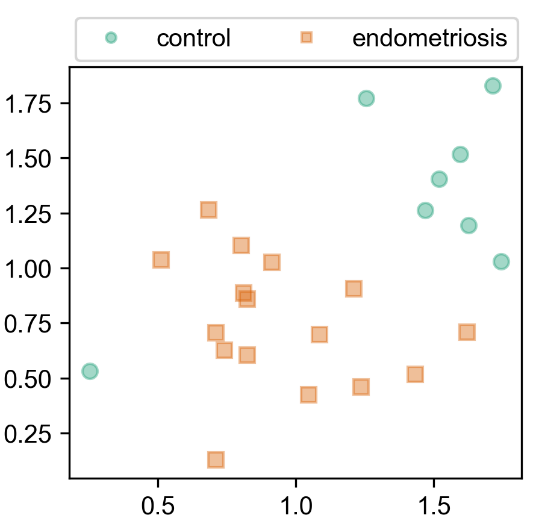

**Supplementary Figure 12.** Classification performance and expression of selected transcripts in all the participants from the secretory group Left: Receiver operating characteristic (ROC) curve showing the predictive performance of the SVM model for all participants in the secretory group. The model was trained using the set of transcripts identified through the feature-selection procedure initiated with random forest importance. Right: Scatter plot of TPM values of transcripts from transcript model 2 from all participants in the secretory group.

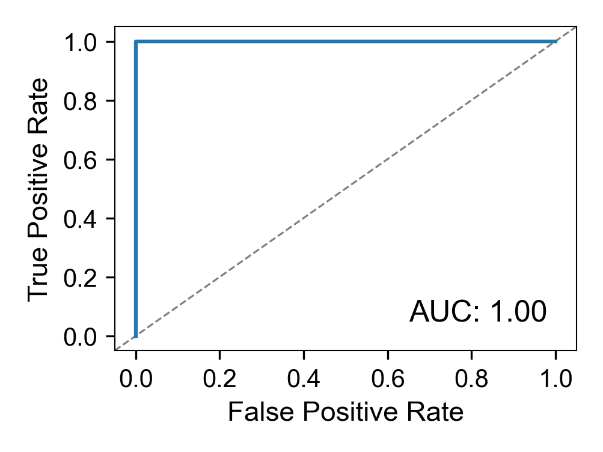

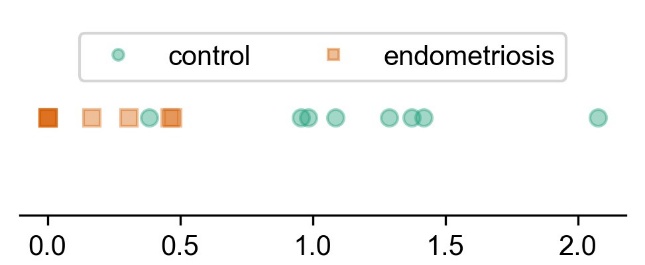

**Supplementary Figure 13.** Classification performance and expression of transcript from transcript model 3 between controls and cases with peritoneal endometriosis in the secretory group. Left: Receiver operating characteristic (ROC) curve showing the predictive performance of the SVM model distinguishing controls from cases with peritoneal endometriosis only in the secretory group. The model was trained using the transcript from transcript model 3, identified through the feature-selection procedure initiated with SVM weights. Right: Plot of TPM values of transcript from transcript model 3 for the same participants, with points colored according to endometriosis status.

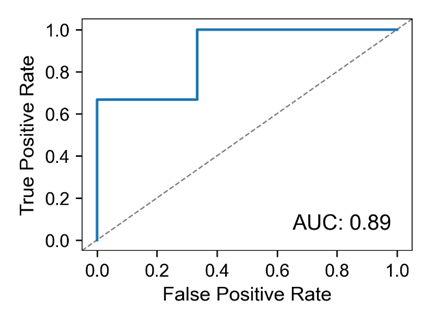

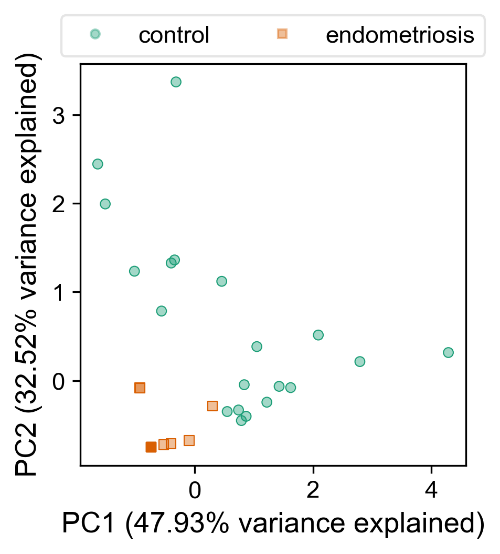

**Supplementary Figure 14**. Classification performance and PCA visualization based on selected transcripts between controls and cases with peritoneal endometriosis in both proliferative and secretory group. Left: Receiver operating characteristic (ROC) curve showing the predictive performance of the SVM model distinguishing controls from cases with peritoneal endometriosis only. The model was trained using the set of transcripts identified through the feature-selection procedure initiated with mutual information. Right: Principal component analysis (PCA) plot generated using the same selected transcript set for participants without endometriosis and those with peritoneal endometriosis only.

References: 1.	Oliveros JC. enny. An interactive tool for comparing lists with Venn's diagrams. Accessed 23.07.2025, <https://bioinfogp.cnb.csic.es/tools/venny/index.html>
