## Supplementary Material_Tables for "Transcriptome profiling to identify blood biomarkers for peritoneal endometriosis"

**Supplementary Tables**

**Supplementary Table 1.** Top 30 enriched pathways for the downregulated genes between the following groups: endometriosis vs controls, peritoneal endometriosis vs controls, peritoneal + ovarian endometriosis vs controls, peritoneal vs peritoneal + ovarian endometriosis in the secretory group. Pathway enrichment was performed using g:Profiler. Pathways are ranked by statistical significance (adjusted p-value < 0.05). Each entry shows the source database, pathway name and ID, adjusted p-value, the number of differentially expressed genes (DGEs) contributing to the enrichment, and the list of intersecting genes.

| **Database** | **Term name** | **Term ID** | **Adjusted p-value** | **Number of intersection genes** | **Intersection genes** |
| --- | --- | --- | --- | --- | --- |
| **ENDOMETRIOSIS vs CONTROLS** | | | | | |
| GO:MF | galactosylgalactosylxylosylprotein 3-beta-glucuronosyltransferase activity | GO:0015018 | 3,31E-02 | 1 | B3GAT1 |
| **PERITONEAL vs CONTROLS** | | | | | |
| GO:MF | cytoskeletal motor activity | GO:0003774 | 4,01E-02 | 8 | KIF19,MYO1E,STARD9,MYO3B,MYO6,MYO19,DYNC1H1,DNHD1 |
| GO:BP | CD4-positive, alpha-beta T cell activation | GO:0035710 | 8,79E-03 | 12 | RUNX3,SH3RF1,KMT2A,CBLB,CD3E,SMAD7,KLHL25,RORA,NKG7,PDP2,IFNG,SLAMF6 |
| GO:BP | T cell differentiation | GO:0030217 | 1,40E-02 | 19 | RUNX3,SH3RF1,KMT2A,PHF10,CD3E,IKZF3,CDK6,SMAD7,KLHL25,RORA,TRAF3IP2,SOS1,ZAP70,WNT10B,ZBTB1,PDP2,IFNG,TCF7,SLAMF6 |
| GO:BP | T cell activation | GO:0042110 | 2,44E-02 | 27 | RUNX3,SH3RF1,KMT2A,PHF10,SMAD3,CBLB,CD3E,IKZF3,CDK6,NLRC3,SMAD7,KLHL25,RORA,TRAF3IP2,NKG7,SOS1,ZAP70,CD40LG,WNT10B,ZBTB1,PDP2,IFNG,TCF7,CD84,SLAMF6,PLXNA1,ITGAL |
| **PERITONEAL + OVARIAN vs CONTROLS** | | | | | |
| KEGG | Mannose type O-glycan biosynthesis | KEGG:00515 | 4,42E-02 | 1 | B3GAT1 |
| **PERITONEAL vs PERITONEAL + OVARIAN** | | | | | |
| GO:BP | killing by host of symbiont cells | GO:0051873 | 2,45E-02 | 2 | CAMP,ARG1 |
| GO:BP | innate immune response | GO:0045087 | 2,99E-02 | 5 | CAMP,CRISP3,ARG1,KRT1,OASL |
| GO:BP | biological process involved in interaction with symbiont | GO:0051702 | 3,55E-02 | 2 | CAMP,ARG1 |
| GO:BP | defense response to symbiont | GO:0140546 | 4,73E-02 | 5 | CAMP,CRISP3,ARG1,KRT1,OASL |
| GO:CC | specific granule lumen | GO:0035580 | 2,13E-05 | 3 | CAMP,CRISP3,ARG1 |
| GO:CC | specific granule | GO:0042581 | 3,79E-04 | 3 | CAMP,CRISP3,ARG1 |
| GO:CC | tertiary granule lumen | GO:1904724 | 1,95E-03 | 2 | CAMP,CRISP3 |
| GO:CC | secretory granule lumen | GO:0034774 | 3,23E-03 | 3 | CAMP,CRISP3,ARG1 |
| GO:CC | cytoplasmic vesicle lumen | GO:0060205 | 3,38E-03 | 3 | CAMP,CRISP3,ARG1 |
| GO:CC | vesicle lumen | GO:0031983 | 3,42E-03 | 3 | CAMP,CRISP3,ARG1 |
| GO:CC | tertiary granule | GO:0070820 | 1,73E-02 | 2 | CAMP,CRISP3 |
| GO:CC | secretory granule | GO:0030141 | 1,84E-02 | 4 | CAMP,CRISP3,ARG1,KRT1 |
| GO:CC | extracellular space | GO:0005615 | 3,40E-02 | 7 | TACSTD2,CAMP,CRISP3,ARG1,KRT1,RHOB,GALM |
| GO:CC | secretory vesicle | GO:0099503 | 3,86E-02 | 4 | CAMP,CRISP3,ARG1,KRT1 |
| REAC | neutrophil degranulation | REAC:R-HSA-6798695 | 4,74E-02 | 4 | CAMP,CRISP3,ARG1,KRT1 |

**Supplementary table 2.** Top 30 enriched pathways for the upregulated genes between the following groups: endometriosis vs controls, peritoneal endometriosis vs controls, peritoneal + ovarian endometriosis vs controls, peritoneal vs peritoneal + ovarian endometriosis in the secretory group. Pathway enrichment was performed using g:Profiler. Pathways are ranked by statistical significance (adjusted p-value < 0.05). Each entry shows the source database, pathway name and ID, adjusted p-value, the number of differentially expressed genes (DGEs) contributing to the enrichment, and the list of intersecting genes.

| **Database** | **Term name** | **Term ID** | **Adjusted p-value** | **Number of intersection genes** | **Intersection genes** |
| --- | --- | --- | --- | --- | --- |
| **ENDOMETRIOSIS vs CONTROLS** | | | | | |
| GO:MF | lipopolysaccharide binding | GO:0001530 | 5,42E-03 | **3** | NINJ1,CD14,PTAFR |
| GO:MF | lipopolysaccharide immune receptor activity | GO:0001875 | 7,45E-03 | **2** | CD14,PTAFR |
| GO:BP | angiogenesis | GO:0001525 | 2,74E-02 | **3** | CD93,CXCL8,NINJ1 |
| GO:BP | blood vessel morphogenesis | GO:0048514 | 4,28E-02 | **3** | CD93,CXCL8,NINJ1 |
| TF | Factor: P53 |  |  |  |  |
| CORUM | NINJ1 homo-oligomer complex | CORUM:6466 | 5,00E-02 | 1 | NINJ1 |
| **PERITONEAL vs CONTROLS** | | | | | |
| GO:MF | lipid binding | GO:0008289 | 6,58E+15 | 52 | FUZ,MPPE1,MAP1LC3A,CAMP,NCF1,NINJ1,CD14,CPNE2,PTAFR,ACOX1,ALOX5AP,SGK1,RXRA,S100A9,LTF,UNC119,GRAMD1C,ABCA1,BAD,MME,NCF4,PLD1,FES,RAPGEF2,ABCG1,S1PR4,BPI,PACSIN2,FCGR3B,OSBPL2,GABARAP,CD300A,RBP7,S100A8,TLR2,CLEC4E,CHMP3,PTEN,CD55,GABARAPL1,NLRP3,RAB35,PICALM,RCSD1,SDCBP,CHMP2A,RUBCNL,RARA,AKT1,SNX10,JAG1,IGF2R |
| GO:CC | cytoplasmic vesicle | GO:0031410 | 1,08E-15 | 180 | RHOB,LAMTOR4,CFP,MAP1LC3A,CAMP,CRISP3,NCF1,CHMP1B,KRT1,CD93,CD14,CPPED1,MNDA,CHIC2,MEFV,PTAFR,ATP6V0B,FGL2,CORO1A,STX10,RAB11FIP1,S100A9,GMFG,RAB31,CEACAM8,TYROBP,ARG1,LTF,IFNGR2,CTSS,SYK,TASL,DYNC1LI1,ANKRD13A,ATP6V1B2,FCGRT,PRDX5,PSEN1,TCN1,UNC93B1,ABCA1,HLA-C,ATP6V0D1,TUBA1A,PYCARD,CHMP5,MME,UBC,APAF1,HSPA6,CHIT1,RHOG,NCF2,CTSZ,ABCA13,NCF4,PRCP,PLD1,SLC31A2,WASHC1,ARRDC3,FES,RAPGEF2,MAPK3,RNF13,OPRL1,FPR1,GRB2,SLC15A3,ITM2B,ABCG1,YPEL5,IRAG2,QPCT,CD82,CLEC5A,ITGA5,DENND10,CRISPLD2,APLP2,BPI,ALOX5,ARPC5,HVCN1,GNAI2,PACSIN2,PIP4P2,FCGR3B,SLC66A2,LAMTOR1,CYBA,FPR2,CDC42,SNAP23,HEBP2,PHF24,PLIN3,RHOA,FTL,OLFM4,GABARAP,LYZ,CD300A,MMP8,LAMP2,TMEM59,CTNNA1,AGPAT2,SRGN,S100A8,TLR2,LRG1,BRI3,RAC1,DNAJC5,CXCL1,CLEC4E,CHMP3,SPACA6,ATP6V1A,CMTM6,BST1,DDIT3,RAB13,RAB5A,CD63,CD55,ERGIC1,PSENEN,GABARAPL1,ARRB2,RGS19,MMP9,DYNLT1,IRF7,F2RL1,RAB35,PICALM,MOSPD2,CXCR1,RCSD1,RP2,RCBTB2,TRAPPC14,SDCBP,HSPA1A,LRRK2,CHMP2A,GM2A,FLOT1,RUBCNL,SPAG9,ANTXR2,TRAPPC3,RAB18,OSCAR,RNF149,AP3S1,NEU1,SCARF1,RNASE6,SNX10,CLTCL1,IGF2R,CLEC4D,CD58,BICD2,SLC15A4,SPAST,IFITM2,ACAP2,FCN1,USP10,PLAUR,RETN,COPE,GOLGA2,DEGS1,ATP6V0E1,OSTF1 |
| GO:CC | intracellular vesicle | GO:0097708 | 1,46E-15 | 180 | RHOB,LAMTOR4,CFP,MAP1LC3A,CAMP,CRISP3,NCF1,CHMP1B,KRT1,CD93,CD14,CPPED1,MNDA,CHIC2,MEFV,PTAFR,ATP6V0B,FGL2,CORO1A,STX10,RAB11FIP1,S100A9,GMFG,RAB31,CEACAM8,TYROBP,ARG1,LTF,IFNGR2,CTSS,SYK,TASL,DYNC1LI1,ANKRD13A,ATP6V1B2,FCGRT,PRDX5,PSEN1,TCN1,UNC93B1,ABCA1,HLA-C,ATP6V0D1,TUBA1A,PYCARD,CHMP5,MME,UBC,APAF1,HSPA6,CHIT1,RHOG,NCF2,CTSZ,ABCA13,NCF4,PRCP,PLD1,SLC31A2,WASHC1,ARRDC3,FES,RAPGEF2,MAPK3,RNF13,OPRL1,FPR1,GRB2,SLC15A3,ITM2B,ABCG1,YPEL5,IRAG2,QPCT,CD82,CLEC5A,ITGA5,DENND10,CRISPLD2,APLP2,BPI,ALOX5,ARPC5,HVCN1,GNAI2,PACSIN2,PIP4P2,FCGR3B,SLC66A2,LAMTOR1,CYBA,FPR2,CDC42,SNAP23,HEBP2,PHF24,PLIN3,RHOA,FTL,OLFM4,GABARAP,LYZ,CD300A,MMP8,LAMP2,TMEM59,CTNNA1,AGPAT2,SRGN,S100A8,TLR2,LRG1,BRI3,RAC1,DNAJC5,CXCL1,CLEC4E,CHMP3,SPACA6,ATP6V1A,CMTM6,BST1,DDIT3,RAB13,RAB5A,CD63,CD55,ERGIC1,PSENEN,GABARAPL1,ARRB2,RGS19,MMP9,DYNLT1,IRF7,F2RL1,RAB35,PICALM,MOSPD2,CXCR1,RCSD1,RP2,RCBTB2,TRAPPC14,SDCBP,HSPA1A,LRRK2,CHMP2A,GM2A,FLOT1,RUBCNL,SPAG9,ANTXR2,TRAPPC3,RAB18,OSCAR,RNF149,AP3S1,NEU1,SCARF1,RNASE6,SNX10,CLTCL1,IGF2R,CLEC4D,CD58,BICD2,SLC15A4,SPAST,IFITM2,ACAP2,FCN1,USP10,PLAUR,RETN,COPE,GOLGA2,DEGS1,ATP6V0E1,OSTF1 |
| GO:CC | secretory granule | GO:0030141 | 2,18E-14 | 98 | CFP,CAMP,CRISP3,KRT1,CD93,CD14,CPPED1,MNDA,PTAFR,FGL2,S100A9,GMFG,RAB31,CEACAM8,TYROBP,ARG1,LTF,CTSS,DYNC1LI1,PSEN1,TCN1,HLA-C,PYCARD,MME,APAF1,HSPA6,CHIT1,RHOG,NCF2,CTSZ,ABCA13,PRCP,PLD1,FPR1,YPEL5,IRAG2,QPCT,CLEC5A,CRISPLD2,APLP2,BPI,ALOX5,ARPC5,HVCN1,GNAI2,FCGR3B,LAMTOR1,CYBA,FPR2,CDC42,SNAP23,HEBP2,RHOA,FTL,OLFM4,LYZ,CD300A,MMP8,LAMP2,CTNNA1,AGPAT2,SRGN,S100A8,TLR2,LRG1,BRI3,RAC1,DNAJC5,CXCL1,SPACA6,ATP6V1A,CMTM6,BST1,RAB13,CD63,CD55,MMP9,DYNLT1,MOSPD2,CXCR1,RCBTB2,SDCBP,HSPA1A,GM2A,SPAG9,RAB18,OSCAR,NEU1,SNX10,IGF2R,CLEC4D,CD58,SLC15A4,FCN1,PLAUR,RETN,DEGS1,OSTF1 |
| GO:CC | secretory vesicle | GO:0099503 | 9,16E-13 | 106 | CFP,CAMP,CRISP3,KRT1,CD93,CD14,CPPED1,MNDA,PTAFR,FGL2,STX10,S100A9,GMFG,RAB31,CEACAM8,TYROBP,ARG1,LTF,CTSS,DYNC1LI1,ATP6V1B2,PSEN1,TCN1,HLA-C,ATP6V0D1,PYCARD,MME,APAF1,HSPA6,CHIT1,RHOG,NCF2,CTSZ,ABCA13,PRCP,PLD1,FPR1,YPEL5,IRAG2,QPCT,CLEC5A,CRISPLD2,APLP2,BPI,ALOX5,ARPC5,HVCN1,GNAI2,FCGR3B,LAMTOR1,CYBA,FPR2,CDC42,SNAP23,HEBP2,PHF24,RHOA,FTL,OLFM4,LYZ,CD300A,MMP8,LAMP2,CTNNA1,AGPAT2,SRGN,S100A8,TLR2,LRG1,BRI3,RAC1,DNAJC5,CXCL1,SPACA6,ATP6V1A,CMTM6,BST1,RAB13,RAB5A,CD63,CD55,MMP9,DYNLT1,PICALM,MOSPD2,CXCR1,RCBTB2,SDCBP,HSPA1A,LRRK2,GM2A,SPAG9,RAB18,OSCAR,AP3S1,NEU1,SNX10,IGF2R,CLEC4D,CD58,SLC15A4,FCN1,PLAUR,RETN,DEGS1,OSTF1 |
| GO:CC | vesicle | GO:0031982 | 6,92E-12 | 227 | FUZ,RHOB,LAMTOR4,TACSTD2,CFP,MAP1LC3A,CAMP,CRISP3,NCF1,B3GNT8,CHMP1B,KRT1,CD93,CD14,CPPED1,MNDA,CHIC2,CPNE2,MEFV,PTAFR,ATP6V0B,FGL2,CORO1A,STX10,LAT2,RAB11FIP1,VASP,G6PD,S100A9,GMFG,RAB31,CEACAM8,ANGPT1,H3-3A,TYROBP,SPINT1,ARG1,EPHB4,LTF,IFNGR2,GLIPR2,CTSS,SYK,TASL,DYNC1LI1,ANKRD13A,ATP6V1B2,LSP1,TALDO1,FCGRT,TAGLN2,PRDX5,ICAM3,PSEN1,TCN1,UNC93B1,ABCA1,HLA-C,ATP6V0D1,TUBA1A,PYCARD,CHMP5,MME,UBC,APAF1,HSPA6,CHIT1,GNB4,RHOG,KCNE3,NCF2,CTSZ,ABCA13,NCF4,PRCP,PLD1,SLC31A2,WASHC1,ARRDC3,FES,RAPGEF2,MAPK3,RNF13,OPRL1,FPR1,GRB2,SLC15A3,ITM2B,ABCG1,YPEL5,IRAG2,QPCT,CD82,CLEC5A,ITGA5,DENND10,CALM2,DDAH2,CRISPLD2,APLP2,FCGR3A,BPI,ALOX5,ARPC5,HVCN1,GNAI2,PACSIN2,PIP4P2,FCGR3B,SLC66A2,LAMTOR1,CYBA,FPR2,PSMB3,H3-3B,CLIC1,CDC42,SNAP23,HEBP2,PHF24,PLIN3,RHOA,FTL,ARPC3,OLFM4,GABARAP,LYZ,CD300A,MMP8,LAMP2,TMEM59,TNFSF10,PLOD1,DAAM2,CTNNA1,AGPAT2,S100A4,SRGN,TYK2,TNFRSF8,S100A8,TLR2,LRG1,BRI3,RAC1,DNAJC5,CXCL1,CLEC4E,CHMP3,SPACA6,ATP6V1A,CMTM6,BST1,DDIT3,HNRNPK,RAB13,RAB5A,CD63,ST3GAL6,CD55,IL1RN,ERGIC1,C11ORF54,PSENEN,GABARAPL1,ARRB2,RGS19,TKT,MMP9,DYNLT1,IRF7,F2RL1,RAB35,S100A6,PICALM,TXN,MOSPD2,CXCR1,ARPC4,RCSD1,RP2,RCBTB2,TRAPPC14,SDCBP,HSPA1A,LRRK2,CHMP2A,GM2A,PTTG1IP,FLOT1,RUBCNL,SPAG9,ANTXR2,TRAPPC3,RAB18,OSCAR,RNF149,AP3S1,NEU1,SCARF1,SOD2,AKT1,RNASE6,SNX10,CLTCL1,ARPC1B,IGF2R,CLEC4D,ALDH2,CD58,BICD2,CYBRD1,SLC15A4,SPAST,SPP1,IFITM2,ACAP2,FCN1,CAPZA2,USP10,PLAUR,RETN,COPE,GOLGA2,DEGS1,ATP6V0E1,OSTF1 |
| GO:CC | endomembrane system | GO:0012505 | 1,25E-08 | 251 | SULF2,RHOB,LAMTOR4,CFP,MPPE1,MAP1LC3A,CAMP,CRISP3,IKBIP,B3GNT8,CHMP1B,GPAT3,KRT1,PIGX,CD93,CD14,CPPED1,MNDA,ST6GALNAC2,CHIC2,PTAFR,ATP6V0B,RASSF2,ALOX5AP,FGL2,CORO1A,STX10,SGK1,DHRS9,RAB11FIP1,S100A9,GMFG,RAB31,CEACAM8,TYROBP,ARG1,NUP214,LTF,IFNGR2,GLIPR2,CTSS,GRAMD1C,TASL,MANSC1,DYNC1LI1,HTATIP2,ANKRD13A,ATP6V1B2,FCGRT,CYP4F3,TMBIM4,PSEN1,TCN1,HMGCR,UNC93B1,ABCA1,HLA-C,ATP6V0D1,TUBA1A,CARD19,PYCARD,CHMP5,MME,UBC,APAF1,LST1,HSPA6,CHIT1,SERINC1,RHOG,GRAMD4,NCF2,CTSZ,ABCA13,PAK1,NCF4,SLC16A3,PRCP,PLD1,SLC31A2,WASHC1,ARRDC3,FES,DHRS7B,RAPGEF2,MAPK3,RNF13,FPR1,GRB2,TRIQK,SLC15A3,ITM2B,ABCG1,RAB5IF,YPEL5,TBXAS1,NHS,IRAG2,FUT7,SFT2D1,VMP1,QPCT,LBR,CLEC5A,ITGA5,DENND10,CRISPLD2,APLP2,BPI,ALOX5,ARPC5,HVCN1,GNAI2,NT5C3A,PACSIN2,PIP4P2,FCGR3B,SLC66A2,LAMTOR1,CYBA,FPR2,OSBPL2,CLIC1,TXNDC12,CDC42,SNAP23,HEBP2,PHF24,PLIN3,RHOA,TMEM43,FTL,OLFM4,CHST15,GABARAP,LYZ,CD300A,MMP8,LAMP2,TMEM59,PLOD1,CTNNA1,AGPAT2,SRGN,FOS,S100A8,TLR2,PISD,LRG1,TMEM120A,TMX4,USF1,BRI3,RAC1,RAF1,DNAJC5,CXCL1,CHMP3,SPACA6,IFIT2,ZDHHC7,ATP6V1A,CMTM6,RNF41,BST1,DDIT3,RAB13,C6ORF89,RAB5A,CD63,ST3GAL6,CHST7,CD55,ERGIC1,PSENEN,GABARAPL1,NLRP3,ARRB2,RSAD2,RGS19,TKT,GAPT,SDC2,MMP9,DYNLT1,IRF7,F2RL1,RAB35,S100A6,PICALM,CCDC88B,MOSPD2,RETREG2,CXCR1,ADAM17,RCSD1,RP2,HSD17B13,SDF2,RCBTB2,TRAPPC14,ANP32A,SDCBP,HSPA1A,TMCC3,FAM114A1,EXTL3,CYB5R4,GALNT2,LRRK2,CHMP2A,HSD17B11,LRRC25,GM2A,FLOT1,TMCC1,PTGES,SPAG9,ANTXR2,TRAPPC3,RAB18,EDEM2,OSCAR,RNF149,AP3S1,NEU1,MTMR6,SNX10,CLTCL1,PIGB,IGF2R,CLEC4D,CD58,BICD2,MX2,SLC15A4,SPAST,SPP1,IFITM2,DNAJB12,ACAP2,FCN1,USP10,PLAUR,RETN,COPE,USP19,GOLGA2,DEGS1,ATP6V0E1,OSTF1 |
| GO:CC | tertiary granule | GO:0070820 | 6,38E-06 | 35 | CFP,CAMP,CRISP3,CD93,PTAFR,CEACAM8,LTF,CTSS,DYNC1LI1,TCN1,CHIT1,PRCP,PLD1,FPR1,YPEL5,QPCT,CLEC5A,LAMTOR1,CYBA,FPR2,SNAP23,RHOA,OLFM4,LYZ,CD300A,MMP8,LAMP2,LRG1,RAC1,CXCL1,CD55,MMP9,OSCAR,CLEC4D,CD58 |
| REAC | Neutrophil degranulation | REAC:R-HSA-6798695 | 1,17E-04 | 86 | CFP,CAMP,CRISP3,KRT1,CD93,CD14,CPPED1,MNDA,PTAFR,FGL2,S100A9,GMFG,RAB31,CEACAM8,TYROBP,ARG1,LTF,CTSS,DYNC1LI1,PSEN1,TCN1,HLA-C,PYCARD,MME,APAF1,HSPA6,CHIT1,RHOG,CTSZ,ABCA13,PRCP,PLD1,FPR1,YPEL5,IRAG2,QPCT,CLEC5A,CRISPLD2,BPI,ALOX5,ARPC5,HVCN1,FCGR3B,LAMTOR1,CYBA,FPR2,SNAP23,HEBP2,RHOA,FTL,OLFM4,LYZ,CD300A,MMP8,LAMP2,AGPAT2,S100A8,TLR2,LRG1,BRI3,RAC1,DNAJC5,CXCL1,CMTM6,BST1,CD63,CD55,MMP9,DYNLT1,MOSPD2,CXCR1,SDCBP,HSPA1A,GM2A,RAB18,OSCAR,NEU1,IGF2R,CLEC4D,CD58,SLC15A4,FCN1,PLAUR,RETN,DEGS1,OSTF1 |
| GO:CC | specific granule | GO:0042581 | 1,60E-04 | 36 | CFP,CAMP,CRISP3,CD93,CEACAM8,ARG1,LTF,TCN1,CHIT1,CTSZ,PLD1,QPCT,CLEC5A,BPI,HVCN1,LAMTOR1,CYBA,FPR2,SNAP23,OLFM4,LYZ,MMP8,AGPAT2,LRG1,DNAJC5,CXCL1,CMTM6,BST1,MOSPD2,OSCAR,NEU1,CLEC4D,SLC15A4,PLAUR,RETN,DEGS1 |
| GO:BP | defense response | GO:0006952 | 5,08E-04 | 129 | CFP,CAMP,CRISP3,CXCL8,NCF1,KRT1,NINJ1,CD14,MNDA,MEFV,PTAFR,ALOX5AP,FGL2,CORO1A,THEMIS2,TSPAN2,S100A9,TMIGD3,TYROBP,C5AR2,ARG1,PVR,LTF,IFNGR2,CTSS,RNF166,SYK,PBXIP1,TASL,LSP1,MSRB1,PRDX5,PELI1,PSEN1,CYSLTR1,UNC93B1,TREM1,HLA-C,LILRA5,OASL,PYCARD,CCN3,HMGB2,CXCL5,GRAMD4,NCF2,NT5C2,PAK1,PRCP,DHRS7B,MAPK3,FPR1,GRB2,SLC15A3,LEP,FUT7,CLEC5A,IL10RB,IFNGR1,FCGR3A,BPI,ALOX5,HVCN1,IL6R,NT5C3A,FCGR3B,CYBA,FPR2,IER3,CDC42,SNAP23,TMEM43,LYZ,CD300A,MMP8,LAMP2,LXN,FOS,TYK2,S100A8,POLB,CASP8,CLEC7A,LILRA2,TLR2,RAC1,RAF1,PADI4,AIF1,CXCL1,CLEC4E,CHMP3,IFIT2,BST1,IFIT3,TXNIP,CD55,IL1RN,ISG20,IL1RAP,NLRP3,ARRB2,RSAD2,MMP9,ZNFX1,IRF7,F2RL1,CCDC88B,ADAM17,HSPA1A,EXTL3,LRRK2,STAT6,FLOT1,PTGES,TRAFD1,AKT1,RNASE6,FADD,CLEC4D,CD58,LRCH4,MX2,RNASE4,SLC15A4,TNFAIP8L2,ISG15,IFITM2,FCN1 |
| GO:CC | bounding membrane of organelle | GO:0098588 | 1,06E-03 | 143 | RHOB,LAMTOR4,CFP,MPPE1,MAP1LC3A,B3GNT8,CHMP1B,CD93,CD14,BID,ST6GALNAC2,FAR2,PTAFR,ACOX1,ATP6V0B,TM6SF1,CORO1A,STX10,RAB11FIP1,RAB31,CEACAM8,TYROBP,BBC3,MTX1,IFNGR2,GLIPR2,PRRG4,TASL,DYNC1LI1,ATP6V1B2,FCGRT,PIGBOS1,TMBIM4,PSEN1,HMGCR,UNC93B1,BAD,HLA-C,ATP6V0D1,BLOC1S2,PYCARD,CHMP5,MME,UBC,LST1,GNB4,RHOG,CTSZ,ABCA13,NCF4,PRCP,PLD1,SLC31A2,WASHC1,DHRS7B,RNF13,FPR1,SLC15A3,ITM2B,ABCG1,SNN,IRAG2,FUT7,VMP1,CLEC5A,APLP2,HVCN1,PACSIN2,PIP4P2,FCGR3B,LAMTOR1,CYBA,FPR2,MCL1,CDC42,SNAP23,PHF24,PLIN3,RHOA,CHST15,GABARAP,CD300A,LAMP2,TMEM59,PLOD1,AGPAT2,CASP8,TLR2,BRI3,RAC1,RAF1,DNAJC5,CLEC4E,CHMP3,SPACA6,ZDHHC7,ATP6V1A,CMTM6,BST1,RAB13,C6ORF89,RAB5A,CD63,ST3GAL6,CHST7,CD55,ERGIC1,PSENEN,GABARAPL1,NLRP3,ARRB2,RSAD2,ZNFX1,IRF7,RAB35,MOSPD2,CXCR1,ADAM17,GALNT2,LRRK2,CHMP2A,FLOT1,RUBCNL,SPAG9,ANTXR2,TRAPPC3,RAB18,NEU1,SCARF1,SNX10,CLTCL1,IGF2R,CLEC4D,CD58,CYBRD1,SLC15A4,IFITM2,ACAP2,PLAUR,COPE,GOLGA2,DEGS1,ATP6V0E1 |
| GO:CC | secretory granule membrane | GO:0030667 | 7,91E-03 | 46 | CD93,CD14,PTAFR,RAB31,CEACAM8,TYROBP,DYNC1LI1,PSEN1,HLA-C,MME,RHOG,ABCA13,PRCP,PLD1,FPR1,IRAG2,CLEC5A,APLP2,HVCN1,FCGR3B,LAMTOR1,CYBA,FPR2,SNAP23,RHOA,CD300A,LAMP2,AGPAT2,TLR2,BRI3,RAC1,DNAJC5,SPACA6,CMTM6,BST1,CD63,CD55,MOSPD2,CXCR1,RAB18,IGF2R,CLEC4D,CD58,SLC15A4,PLAUR,DEGS1 |
| GO:BP | response to external stimulus | GO:0009605 | 2,80E-02 | 150 | CFP,CAMP,CRISP3,CXCL8,NCF1,KRT1,NINJ1,CD14,MNDA,MEFV,PTAFR,ALOX5AP,FGL2,CORO1A,S100A9,ANGPT1,TYROBP,C5AR2,ARG1,PVR,LTF,IFNGR2,CTSS,UNC119,RNF166,SYK,CMTM2,TASL,LSP1,MSRB1,PELI1,CYSLTR1,UNC93B1,ABCA1,TREM1,BAD,HLA-C,TUBA1A,LILRA5,OASL,PYCARD,CHMP5,CCN3,CHIT1,HMGB2,RHOG,CKLF,CXCL5,GRAMD4,NCF2,NT5C2,PAK1,PLD1,FES,MAPK3,FPR1,GRB2,SLC15A3,LEP,FUT7,CLEC5A,IL10RB,IFNGR1,FCGR3A,BPI,ALOX5,HVCN1,IL6R,NT5C3A,CYBA,FPR2,IER3,CDC42,PHF24,RHOA,TMEM43,LYZ,CD300A,MMP8,LAMP2,LXN,S100A4,FOS,TYK2,TNFRSF8,S100A8,CASP8,CLEC7A,LILRA2,TLR2,LRG1,TMEM120A,USF1,RAC1,RAF1,TYMP,PADI4,AIF1,CXCL1,CLEC4E,MAP2K4,CHMP3,IFIT2,PTEN,BST1,RAB13,IFIT3,TXNIP,CD63,CD55,ISG20,IL1RAP,NLRP3,ARRB2,RSAD2,MMP9,ZNFX1,IRF7,F2RL1,CCDC88B,MOSPD2,CXCR1,ADAM17,HSPA1A,EXTL3,LRRK2,FLOT1,PTGES,RARA,TRAFD1,XPC,SCARF1,SOD2,AKT1,RNASE6,FADD,CLEC4D,CD58,LRCH4,MX2,RNASE4,SLC15A4,TNFSF14,TNFAIP8L2,SPP1,ISG15,IFITM2,FCN1,PLAUR,RETN |
| GO:CC | cytoplasm | GO:0005737 | 7,99E-02 | 451 | FUZ,SULF2,RHOB,IMPA2,MID1IP1,LAMTOR4,TACSTD2,CFP,MPPE1,MAP1LC3A,CAMP,CRISP3,IKBIP,NCF1,B3GNT8,CHMP1B,GPAT3,KRT1,PIGX,CD93,CD14,CPPED1,MNDA,CFAP45,LETM2,BID,ST6GALNAC2,CHIC2,CPNE2,TNNI2,FAR2,MEFV,JARID2,PTAFR,ACOX1,ADPRS,ATP6V0B,RASSF2,TM6SF1,ALOX5AP,FGL2,CORO1A,STX10,SGK1,DHRS9,FRAT1,RAB11FIP1,THEMIS2,VASP,RXRA,G6PD,S100A9,KAT8,GMFG,RAB31,RILPL2,CEACAM8,TYROBP,SPINT1,ARG1,EPHB4,BBC3,NUP214,MTX1,CEP19,PVR,LTF,IFNGR2,GLIPR2,CTSS,UNC119,GRAMD1C,RNF166,SYK,PBXIP1,RGS2,PRRG4,TASL,CFAP54,MANSC1,DYNC1LI1,HTATIP2,MYCBP,ANKRD13A,TST,ATP6V1B2,TALDO1,FCGRT,MAP7,PELI2,NDUFB6,MSRB1,TAGLN2,PRDX5,CYP4F3,ISCU,KRT23,PELI1,CCDC153,OAZ2,PIGBOS1,TMBIM4,PSEN1,SQOR,MYADM,TCN1,HSH2D,HMGCR,ARID3A,TRIOBP,DAZAP2,UNC93B1,CSTA,ABCA1,BAD,HLA-C,TIMMDC1,ATP6V0D1,B9D2,TUBA1A,OASL,BLOC1S2,CARD19,PYCARD,PPP4R1,CHMP5,MME,CCN3,UBC,APAF1,LST1,HSPA6,CHIT1,SERINC1,HMGB2,GNB4,RHOG,PFKFB4,KCNE3,XKR8,GRAMD4,NCF2,CTSZ,NT5C2,ABCA13,PAK1,NCF4,PANK2,PRCP,PLD1,MLF2,SLC31A2,BACH1,WASHC1,ARRDC3,FES,OPLAH,DHRS7B,RAPGEF2,MAPK3,RNF13,OPRL1,FPR1,GRB2,TRIQK,SLC15A3,ITM2B,ABCG1,LEP,RAB5IF,SNN,YPEL5,TBXAS1,NHS,IRAG2,FUT7,SFT2D1,VMP1,QPCT,S1PR4,CD82,LBR,CLEC5A,RPL3L,ITGA5,DENND10,ITPK1,GLYR1,CALM2,DDAH2,CRISPLD2,APLP2,SAT1,AMACR,BPI,ALOX5,ARPC5,HVCN1,NDC80,GNAI2,ROPN1L,NT5C3A,PACSIN2,PIP4P2,FCGR3B,SLC66A2,LAMTOR1,CYBA,ASF1B,FPR2,OSBPL2,CDKN2B,CD2BP2,PSMB3,CLIC1,TXNDC12,MCL1,SPOP,PLEKHO1,RNF130,IER3,AMD1,PLEKHG3,FRAT2,CDC42,SNAP23,HEBP2,PHF24,PLIN3,ABHD5,RHOA,TMEM43,FTL,EGLN1,ARPC3,OLFM4,CHST15,GABARAP,LYZ,CD300A,MMP8,LAMP2,LXN,TMEM59,RBP7,TATDN3,PLOD1,CTNNA1,AGPAT2,S100A4,SRGN,FOS,HECW2,TYK2,TNFRSF8,S100A8,POLB,CASP8,CLEC7A,TLR2,PISD,LRG1,TMX4,USF1,BRI3,RAC1,ELL,ERF,RAF1,TYMP,PADI4,COQ8A,DNAJC5,AIF1,MSRB2,CXCL1,CLEC4E,DHTKD1,MAP2K4,CHMP3,SPACA6,IFIT2,SETX,TBCC,ZDHHC7,STK38L,FBXL5,ATP6V1A,CMTM6,RNF41,OGFR,TAGAP,PTEN,BST1,PSRC1,DDIT3,MPP7,TAF10,GMPR2,HNRNPK,RAB13,IFIT3,TXNIP,C6ORF89,RAB5A,CD63,ST3GAL6,CHST7,CD55,IL1RN,SH3BP5L,ISG20,CRLF3,ERGIC1,PSENEN,TPD52L2,GABARAPL1,MTHFS,NLRP3,ARRB2,RSAD2,RGS19,TKT,TK2,GAPT,ZFAS1,SDC2,MMP9,ZNFX1,DYNLT1,IRF7,F2RL1,RAB35,S100A6,PICALM,FBXO7,HEXIM2,ENSG00000280571,CCDC88B,TXN,RRM2B,MOSPD2,RETREG2,CXCR1,ARPC4,ADAM17,RCSD1,GTF2I,RP2,HSD17B13,BTBD10,SFXN5,SDF2,ARHGEF40,RCBTB2,TRAPPC14,ANP32A,SDCBP,CCNG2,AGO4,HSPA1A,LYRM1,TMCC3,FAM114A1,COP1,EXTL3,CYB5R4,GALNT2,LRRK2,SRSF8,CHMP2A,NPEPL1,HSD17B11,STAT6,LRRC25,GM2A,PTTG1IP,FLOT1,SHROOM1,ACOT9,TMCC1,CCNY,PPP2R5A,PTGES,RUBCNL,SPAG9,ANTXR2,TRAPPC3,RARA,RAB18,HSDL2,EDEM2,OSCAR,RNF149,TRAFD1,PLBD1,AP3S1,XPC,GNA15,NEU1,KBTBD7,SCARF1,SOD2,AKT1,TXNL4B,RNASE6,MTMR6,SNX10,FADD,CLTCL1,SPOPL,PIGB,TDRD7,ARPC1B,IGF2R,CLEC4D,PPIF,PPP4C,ALDH2,CD58,BICD2,CYBRD1,MX2,SLC15A4,TNFSF14,CDC42SE1,BCKDK,TNFSF13B,TNFAIP8L2,SPAST,SPP1,TUFT1,ISG15,IFITM2,DNAJB12,ACAP2,FCN1,HSPBAP1,CAPZA2,USP10,STK17B,PLAUR,SULT1A1,RETN,COPE,USP19,GOLGA2,DEGS1,ATP6V0E1,OSTF1,NADK |
| GO:BP | defense response to symbiont | GO:0140546 | 5,40E-01 | 88 | CFP,CAMP,CRISP3,CXCL8,NCF1,KRT1,NINJ1,CD14,MNDA,MEFV,CORO1A,S100A9,TYROBP,ARG1,PVR,LTF,IFNGR2,CTSS,RNF166,SYK,TASL,MSRB1,PELI1,UNC93B1,TREM1,HLA-C,LILRA5,OASL,PYCARD,HMGB2,CXCL5,GRAMD4,NCF2,PAK1,GRB2,SLC15A3,LEP,CLEC5A,IL10RB,IFNGR1,FCGR3A,BPI,HVCN1,CYBA,FPR2,CDC42,TMEM43,LYZ,CD300A,LAMP2,TYK2,S100A8,CASP8,CLEC7A,LILRA2,TLR2,RAF1,PADI4,AIF1,CXCL1,CLEC4E,IFIT2,IFIT3,CD55,ISG20,IL1RAP,NLRP3,ARRB2,RSAD2,ZNFX1,IRF7,F2RL1,HSPA1A,FLOT1,TRAFD1,AKT1,RNASE6,FADD,CLEC4D,CD58,LRCH4,MX2,RNASE4,SLC15A4,TNFAIP8L2,ISG15,IFITM2,FCN1 |
| GO:BP | innate immune response | GO:0045087 | 7,09E-01 | 83 | CFP,CAMP,CRISP3,NCF1,KRT1,NINJ1,CD14,MNDA,MEFV,CORO1A,S100A9,TYROBP,ARG1,PVR,LTF,IFNGR2,CTSS,RNF166,SYK,TASL,MSRB1,PELI1,UNC93B1,TREM1,HLA-C,LILRA5,OASL,PYCARD,HMGB2,GRAMD4,NCF2,PAK1,GRB2,SLC15A3,LEP,CLEC5A,IL10RB,IFNGR1,FCGR3A,BPI,HVCN1,CYBA,FPR2,CDC42,TMEM43,CD300A,LAMP2,TYK2,S100A8,CASP8,CLEC7A,LILRA2,TLR2,RAF1,PADI4,AIF1,CLEC4E,IFIT2,IFIT3,CD55,ISG20,IL1RAP,NLRP3,ARRB2,RSAD2,ZNFX1,IRF7,F2RL1,HSPA1A,FLOT1,TRAFD1,AKT1,RNASE6,FADD,CLEC4D,CD58,LRCH4,MX2,SLC15A4,TNFAIP8L2,ISG15,IFITM2,FCN1 |
| GO:BP | response to other organism | GO:0051707 | 1,92E+00 | 108 | CFP,CAMP,CRISP3,CXCL8,NCF1,KRT1,NINJ1,CD14,MNDA,MEFV,PTAFR,FGL2,CORO1A,S100A9,TYROBP,ARG1,PVR,LTF,IFNGR2,CTSS,RNF166,SYK,TASL,MSRB1,PELI1,UNC93B1,ABCA1,TREM1,HLA-C,LILRA5,OASL,PYCARD,CHMP5,CHIT1,HMGB2,CXCL5,GRAMD4,NCF2,NT5C2,PAK1,MAPK3,GRB2,SLC15A3,LEP,CLEC5A,IL10RB,IFNGR1,FCGR3A,BPI,HVCN1,IL6R,NT5C3A,CYBA,FPR2,IER3,CDC42,RHOA,TMEM43,LYZ,CD300A,MMP8,LAMP2,FOS,TYK2,S100A8,CASP8,CLEC7A,LILRA2,TLR2,LRG1,RAF1,PADI4,AIF1,CXCL1,CLEC4E,CHMP3,IFIT2,IFIT3,CD55,ISG20,IL1RAP,NLRP3,ARRB2,RSAD2,MMP9,ZNFX1,IRF7,F2RL1,CCDC88B,ADAM17,HSPA1A,FLOT1,RARA,TRAFD1,SOD2,AKT1,RNASE6,FADD,CLEC4D,CD58,LRCH4,MX2,RNASE4,SLC15A4,TNFAIP8L2,ISG15,IFITM2,FCN1 |
| GO:BP | response to biotic stimulus | GO:0009607 | 5,46E+00 | 111 | CFP,CAMP,CRISP3,CXCL8,NCF1,KRT1,NINJ1,CD14,MNDA,MEFV,PTAFR,FGL2,CORO1A,S100A9,TYROBP,ARG1,PVR,LTF,IFNGR2,CTSS,RNF166,SYK,TASL,MSRB1,PELI1,UNC93B1,ABCA1,TREM1,HLA-C,LILRA5,OASL,PYCARD,CHMP5,APAF1,CHIT1,HMGB2,CXCL5,GRAMD4,NCF2,NT5C2,PAK1,MAPK3,GRB2,SLC15A3,LEP,CLEC5A,IL10RB,IFNGR1,FCGR3A,BPI,HVCN1,IL6R,NT5C3A,CYBA,FPR2,IER3,CDC42,RHOA,TMEM43,LYZ,CD300A,MMP8,LAMP2,FOS,TYK2,S100A8,CASP8,CLEC7A,LILRA2,TLR2,LRG1,RAF1,PADI4,AIF1,CXCL1,CLEC4E,CHMP3,IFIT2,DDIT3,IFIT3,TXNIP,CD55,ISG20,IL1RAP,NLRP3,ARRB2,RSAD2,MMP9,ZNFX1,IRF7,F2RL1,CCDC88B,ADAM17,HSPA1A,FLOT1,RARA,TRAFD1,SOD2,AKT1,RNASE6,FADD,CLEC4D,CD58,LRCH4,MX2,RNASE4,SLC15A4,TNFAIP8L2,ISG15,IFITM2,FCN1 |
| GO:CC | organelle membrane | GO:0031090 | 6,21E+00 | 187 | RHOB,LAMTOR4,CFP,MPPE1,MAP1LC3A,IKBIP,B3GNT8,CHMP1B,GPAT3,PIGX,CD93,CD14,LETM2,BID,ST6GALNAC2,FAR2,PTAFR,ACOX1,ATP6V0B,TM6SF1,ALOX5AP,CORO1A,STX10,SGK1,DHRS9,RAB11FIP1,RAB31,CEACAM8,TYROBP,BBC3,MTX1,IFNGR2,GLIPR2,GRAMD1C,PRRG4,TASL,DYNC1LI1,ATP6V1B2,FCGRT,NDUFB6,CYP4F3,PIGBOS1,TMBIM4,PSEN1,SQOR,HMGCR,UNC93B1,ABCA1,BAD,HLA-C,TIMMDC1,ATP6V0D1,BLOC1S2,CARD19,PYCARD,CHMP5,MME,UBC,LST1,SERINC1,GNB4,RHOG,GRAMD4,CTSZ,ABCA13,PAK1,NCF4,SLC16A3,PRCP,PLD1,SLC31A2,WASHC1,DHRS7B,RNF13,FPR1,GRB2,TRIQK,SLC15A3,ITM2B,ABCG1,RAB5IF,SNN,TBXAS1,IRAG2,FUT7,VMP1,LBR,CLEC5A,APLP2,ALOX5,HVCN1,PACSIN2,PIP4P2,FCGR3B,LAMTOR1,CYBA,FPR2,CLIC1,MCL1,CDC42,SNAP23,PHF24,PLIN3,RHOA,TMEM43,CHST15,GABARAP,CD300A,LAMP2,TMEM59,PLOD1,AGPAT2,CASP8,TLR2,PISD,TMEM120A,TMX4,BRI3,RAC1,RAF1,DNAJC5,CLEC4E,CHMP3,SPACA6,ZDHHC7,ATP6V1A,CMTM6,BST1,RAB13,C6ORF89,RAB5A,CD63,ST3GAL6,CHST7,CD55,ERGIC1,PSENEN,GABARAPL1,NLRP3,ARRB2,RSAD2,TKT,ZNFX1,IRF7,RAB35,MOSPD2,RETREG2,CXCR1,ADAM17,SFXN5,SDCBP,TMCC3,EXTL3,CYB5R4,GALNT2,LRRK2,CHMP2A,FLOT1,TMCC1,PTGES,RUBCNL,SPAG9,ANTXR2,TRAPPC3,RAB18,AP3S1,NEU1,SCARF1,SNX10,CLTCL1,PIGB,IGF2R,CLEC4D,PPIF,CD58,CYBRD1,SLC15A4,SPAST,IFITM2,DNAJB12,ACAP2,PLAUR,COPE,USP19,GOLGA2,DEGS1,ATP6V0E1 |
| GO:BP | defense response to other organism | GO:0098542 | 1,03E+01 | 96 | CFP,CAMP,CRISP3,CXCL8,NCF1,KRT1,NINJ1,CD14,MNDA,MEFV,FGL2,CORO1A,S100A9,TYROBP,ARG1,PVR,LTF,IFNGR2,CTSS,RNF166,SYK,TASL,MSRB1,PELI1,UNC93B1,TREM1,HLA-C,LILRA5,OASL,PYCARD,HMGB2,CXCL5,GRAMD4,NCF2,NT5C2,PAK1,MAPK3,GRB2,SLC15A3,LEP,CLEC5A,IL10RB,IFNGR1,FCGR3A,BPI,HVCN1,IL6R,NT5C3A,CYBA,FPR2,CDC42,TMEM43,LYZ,CD300A,LAMP2,TYK2,S100A8,CASP8,CLEC7A,LILRA2,TLR2,RAF1,PADI4,AIF1,CXCL1,CLEC4E,CHMP3,IFIT2,IFIT3,CD55,ISG20,IL1RAP,NLRP3,ARRB2,RSAD2,ZNFX1,IRF7,F2RL1,CCDC88B,ADAM17,HSPA1A,FLOT1,TRAFD1,AKT1,RNASE6,FADD,CLEC4D,CD58,LRCH4,MX2,RNASE4,SLC15A4,TNFAIP8L2,ISG15,IFITM2,FCN1 |
| GO:CC | vesicle membrane | GO:0012506 | 1,17E+01 | 93 | RHOB,LAMTOR4,CHMP1B,CD93,CD14,PTAFR,ATP6V0B,CORO1A,STX10,RAB11FIP1,RAB31,CEACAM8,TYROBP,IFNGR2,TASL,DYNC1LI1,ATP6V1B2,FCGRT,PSEN1,HLA-C,ATP6V0D1,CHMP5,MME,UBC,RHOG,ABCA13,NCF4,PRCP,PLD1,SLC31A2,WASHC1,RNF13,FPR1,GRB2,SLC15A3,ITM2B,IRAG2,CLEC5A,APLP2,HVCN1,PACSIN2,PIP4P2,FCGR3B,LAMTOR1,CYBA,FPR2,SNAP23,PHF24,PLIN3,RHOA,CD300A,LAMP2,TMEM59,AGPAT2,TLR2,BRI3,RAC1,DNAJC5,CLEC4E,CHMP3,SPACA6,ATP6V1A,CMTM6,BST1,RAB13,RAB5A,CD63,CD55,PSENEN,GABARAPL1,ARRB2,IRF7,RAB35,MOSPD2,CXCR1,LRRK2,CHMP2A,ANTXR2,RAB18,AP3S1,SCARF1,SNX10,CLTCL1,IGF2R,CLEC4D,CD58,SLC15A4,IFITM2,ACAP2,PLAUR,COPE,DEGS1,ATP6V0E1 |
| GO:CC | vacuole | GO:0005773 | 1,24E+01 | 74 | LAMTOR4,MAP1LC3A,NCF1,CHMP1B,CPPED1,MNDA,MEFV,ATP6V0B,TM6SF1,CEACAM8,ARG1,BBC3,CTSS,TASL,ATP6V1B2,PSEN1,UNC93B1,ATP6V0D1,BLOC1S2,PYCARD,CHMP5,CHIT1,GNB4,NCF2,CTSZ,ABCA13,NCF4,PRCP,PLD1,SLC31A2,WASHC1,ARRDC3,RNF13,FPR1,SLC15A3,IRAG2,VMP1,BPI,PIP4P2,LAMTOR1,CDC42,SNAP23,HEBP2,FTL,GABARAP,LYZ,LAMP2,TMEM59,SRGN,BRI3,DNAJC5,CHMP3,ATP6V1A,CMTM6,CD63,GABARAPL1,SDC2,SDCBP,LRRK2,CHMP2A,GM2A,FLOT1,RUBCNL,SPAG9,RAB18,PLBD1,NEU1,RNASE6,CYBRD1,SLC15A4,TNFAIP8L2,IFITM2,RETN,ATP6V0E1 |
| GO:CC | cytoplasmic vesicle membrane | GO:0030659 | 1,38E+01 | 92 | RHOB,LAMTOR4,CHMP1B,CD93,CD14,PTAFR,ATP6V0B,CORO1A,STX10,RAB11FIP1,RAB31,CEACAM8,TYROBP,IFNGR2,TASL,DYNC1LI1,ATP6V1B2,FCGRT,PSEN1,HLA-C,ATP6V0D1,CHMP5,MME,UBC,RHOG,ABCA13,NCF4,PRCP,PLD1,SLC31A2,WASHC1,RNF13,FPR1,SLC15A3,ITM2B,IRAG2,CLEC5A,APLP2,HVCN1,PACSIN2,PIP4P2,FCGR3B,LAMTOR1,CYBA,FPR2,SNAP23,PHF24,PLIN3,RHOA,CD300A,LAMP2,TMEM59,AGPAT2,TLR2,BRI3,RAC1,DNAJC5,CLEC4E,CHMP3,SPACA6,ATP6V1A,CMTM6,BST1,RAB13,RAB5A,CD63,CD55,PSENEN,GABARAPL1,ARRB2,IRF7,RAB35,MOSPD2,CXCR1,LRRK2,CHMP2A,ANTXR2,RAB18,AP3S1,SCARF1,SNX10,CLTCL1,IGF2R,CLEC4D,CD58,SLC15A4,IFITM2,ACAP2,PLAUR,COPE,DEGS1,ATP6V0E1 |
| GO:CC | lysosome | GO:0005764 | 2,09E+01 | 67 | LAMTOR4,MAP1LC3A,NCF1,CHMP1B,CPPED1,MNDA,ATP6V0B,TM6SF1,CEACAM8,ARG1,BBC3,CTSS,TASL,ATP6V1B2,PSEN1,UNC93B1,ATP6V0D1,BLOC1S2,PYCARD,CHMP5,CHIT1,GNB4,NCF2,CTSZ,ABCA13,NCF4,PRCP,PLD1,SLC31A2,ARRDC3,RNF13,FPR1,SLC15A3,IRAG2,BPI,PIP4P2,LAMTOR1,SNAP23,HEBP2,FTL,GABARAP,LYZ,LAMP2,TMEM59,SRGN,BRI3,DNAJC5,CHMP3,ATP6V1A,CMTM6,CD63,SDC2,SDCBP,LRRK2,CHMP2A,GM2A,FLOT1,SPAG9,PLBD1,NEU1,RNASE6,CYBRD1,SLC15A4,TNFAIP8L2,IFITM2,RETN,ATP6V0E1 |
| GO:CC | lytic vacuole | GO:0000323 | 2,09E+01 | 67 | LAMTOR4,MAP1LC3A,NCF1,CHMP1B,CPPED1,MNDA,ATP6V0B,TM6SF1,CEACAM8,ARG1,BBC3,CTSS,TASL,ATP6V1B2,PSEN1,UNC93B1,ATP6V0D1,BLOC1S2,PYCARD,CHMP5,CHIT1,GNB4,NCF2,CTSZ,ABCA13,NCF4,PRCP,PLD1,SLC31A2,ARRDC3,RNF13,FPR1,SLC15A3,IRAG2,BPI,PIP4P2,LAMTOR1,SNAP23,HEBP2,FTL,GABARAP,LYZ,LAMP2,TMEM59,SRGN,BRI3,DNAJC5,CHMP3,ATP6V1A,CMTM6,CD63,SDC2,SDCBP,LRRK2,CHMP2A,GM2A,FLOT1,SPAG9,PLBD1,NEU1,RNASE6,CYBRD1,SLC15A4,TNFAIP8L2,IFITM2,RETN,ATP6V0E1 |
| GO:BP | response to external biotic stimulus | GO:0043207 | 2,20E+01 | 108 | CFP,CAMP,CRISP3,CXCL8,NCF1,KRT1,NINJ1,CD14,MNDA,MEFV,PTAFR,FGL2,CORO1A,S100A9,TYROBP,ARG1,PVR,LTF,IFNGR2,CTSS,RNF166,SYK,TASL,MSRB1,PELI1,UNC93B1,ABCA1,TREM1,HLA-C,LILRA5,OASL,PYCARD,CHMP5,CHIT1,HMGB2,CXCL5,GRAMD4,NCF2,NT5C2,PAK1,MAPK3,GRB2,SLC15A3,LEP,CLEC5A,IL10RB,IFNGR1,FCGR3A,BPI,HVCN1,IL6R,NT5C3A,CYBA,FPR2,IER3,CDC42,RHOA,TMEM43,LYZ,CD300A,MMP8,LAMP2,FOS,TYK2,S100A8,CASP8,CLEC7A,LILRA2,TLR2,LRG1,RAF1,PADI4,AIF1,CXCL1,CLEC4E,CHMP3,IFIT2,IFIT3,CD55,ISG20,IL1RAP,NLRP3,ARRB2,RSAD2,MMP9,ZNFX1,IRF7,F2RL1,CCDC88B,ADAM17,HSPA1A,FLOT1,RARA,TRAFD1,SOD2,AKT1,RNASE6,FADD,CLEC4D,CD58,LRCH4,MX2,RNASE4,SLC15A4,TNFAIP8L2,ISG15,IFITM2,FCN1 |
| REAC | Innate Immune System | REAC:R-HSA-168249 | 8,72E+01 | 127 | CFP,CAMP,CRISP3,IKBIP,NCF1,KRT1,CD93,CD14,CPPED1,MNDA,MEFV,PTAFR,ATP6V0B,FGL2,LAT2,S100A9,GMFG,RAB31,CEACAM8,TYROBP,C5AR2,ARG1,LTF,CTSS,SYK,DYNC1LI1,ATP6V1B2,PELI2,PELI1,ICAM3,PSEN1,TCN1,UNC93B1,TREM1,HLA-C,ATP6V0D1,PYCARD,MME,UBC,APAF1,HSPA6,CHIT1,RHOG,NCF2,CTSZ,ABCA13,PAK1,NCF4,PRCP,PLD1,MAPK3,FPR1,YPEL5,IRAG2,QPCT,CLEC5A,CRISPLD2,FCGR3A,BPI,ALOX5,ARPC5,HVCN1,FCGR3B,LAMTOR1,CYBA,FPR2,PSMB3,CDC42,SNAP23,HEBP2,RHOA,FTL,ARPC3,OLFM4,LYZ,CD300A,MMP8,LAMP2,AGPAT2,FOS,S100A8,CASP8,CLEC7A,TLR2,LRG1,BRI3,RAC1,RAF1,DNAJC5,CXCL1,CLEC4E,MAP2K4,ATP6V1A,CMTM6,BST1,TXNIP,CD63,CD55,MMP9,DYNLT1,IRF7,TXN,MOSPD2,CXCR1,ARPC4,SDCBP,HSPA1A,STAT6,GM2A,RAB18,OSCAR,NEU1,RNASE6,FADD,ARPC1B,IGF2R,CLEC4D,CD58,SLC15A4,ISG15,FCN1,CAPZA2,PLAUR,RETN,DEGS1,ATP6V0E1,OSTF1 |
| **PERITONEAL + OVARIAN vs CONTROLS** | | | | | |
| GO:MF | cysteine dioxygenase activity | GO:0017172 | 2,49E-02 | 1 | CDO1 |
| KEGG | Taurine and hypotaurine metabolism | KEGG:00430 | 3,07E-02 | 1 | CDO1 |
| **PERITONEAL vs PERITONEAL + OVARIAN** | | | | | |
| GO:CC | nuclear proteasome complex | GO:0031595 | 2,06E-02 | 1 | UBQLN4 |
